# Knowledge, attitudes, practices regarding vector-borne diseases among adults in Switzerland: a cross-sectional survey

**DOI:** 10.64898/2026.08.05.26358163

**Authors:** Lilian Goepp, Eva Maria Hodel, Arlette Szelecsenyi, Nina Huber, Ana Maria Vicedo-Cabrera, Ioannis Magouras, Julien Riou

## Abstract

**Background:** Vector-borne diseases (VBDs) are an evolving public health concern in Switzerland, where endemic tick-borne infections coexist with emerging mosquito-borne threats linked to climate and ecological change. Public preparedness depends on population knowledge, risk perception, and preventive behaviour alongside institutional capacity. We assessed knowledge, attitudes, and practices (KAP) regarding VBDs among adults in the canton of Bern, interpreted alongside a complementary national stakeholder survey.

**Methods:** We analysed a 2025 cross-sectional web-based survey embedded in the BEready cohort. After validity screening, we derived a latent knowledge score using a two-parameter logistic item response theory (IRT) model fitted to knowledge items. Multivariable linear regression examined associations between participant characteristics and latent knowledge. We identified KAP profiles through partitioning-around-medoids clustering based on block-weighted Gower dissimilarities. A parallel survey of cantonal and Liechtenstein authorities in human health, animal health, and environment departments provided institutional context.

**Results:** Among 1,847 respondents, 1,337 met validity criteria. Knowledge was strongest for tick-related content: 98% matched tick-borne encephalitis to ticks, 87% did so for Lyme disease. Mosquito-borne knowledge was markedly weaker, with only 36% correctly classifying chikungunya and 43% West Nile fever as mosquito-borne, despite 53% reporting at least weekly summer mosquito exposure. Tick checks were reported by 79% of participants versus 30% for mosquito standing-water removal. The IRT model indicated that mosquito-borne items were both hardest and most discriminating. Higher knowledge was associated with educational attainment, female sex, residence history, tick-bite frequency, and travel history, alongside a non-linear age effect peaking in mid-adulthood. Clustering identified six KAP profiles distinguishing knowledge gaps, low perceived relevance, and weak translation of knowledge into practice. The stakeholder survey (n=55) showed institutional engagement was considerably more developed for mosquito-borne than tick-borne diseases, although about half of authorities reported no dedicated human resources (49%) or budget (55%).

**Conclusions:** Public knowledge and practice remain stronger for tick-borne than mosquito-borne diseases, despite frequent mosquito exposure, revealing a communication gap. Institutional preparedness shows the opposite pattern, being more developed for mosquito-borne threats. Public health strategies should sustain effective tick-prevention messaging while strengthening mosquito-borne disease communication, household source reduction, and support for community-level surveillance and control.

## 1 Background

Vector-borne diseases (VBDs) are an important and evolving public health challenge in Europe, shaped by climatic, ecological, and social changes that affect vector distribution, seasonality, and opportunities for pathogen transmission [1, 2, 3]. In Europe, tick-borne encephalitis (TBE) and Lyme borreliosis remain the predominant endemic VBDs, while risks associated with mosquito-borne diseases are becoming increasingly prominent with the spread of invasive *Aedes* mosquitoes, with recurrent autochthonous transmission events involving arboviruses such as dengue, chikungunya, and West Nile virus [1, 3]. Both dimensions are relevant to Switzerland. Tick-borne diseases remain the dominant recognized VBD threat. Switzerland has one of the highest incidences of Lyme borreliosis in Europe, with an estimated incidence of 130–224 cases per 100,000 person-years ([4, 5, 6]) and a seroprevalence of 6.1% among blood donors ([6]). TBE is a notifiable disease with only 3-5 reported cases per 100,000 person-years, but a recent seroprevalence study in healthcare workers highlighted massive underdetection, with a seroprevalence of 6% among unvaccinated individuals, corresponding to an estimated annual incidence of infection of 735 per 100,000. At the same time, mosquito-borne risks are changing, particularly in relation to the establishment of *Aedes albopictus* in canton Ticino, south of the Alps, since its first detection in 2003 ([7]), and its documented northward spread towards the Swiss plateau ([8, 9]). This raises the prospect of local transmission following travel-related importation of arboviral infections in the near future, in a country where no such events have yet been reported, contrary to neighbouring countries ([10]).

Under these conditions, public health responses to VBDs require not only institutional capacity for surveillance, prevention, and control, but also adequate public awareness, risk perception, and uptake of preventive behaviours. Research on knowledge, attitudes and practices (KAP) related to tick-borne diseases in Europe has shown moderate to high levels of awareness in endemic regions, but that knowledge does not reliably translate into protective behaviour ([11, 12, 13, 14]). Adoption of recommended tick prevention measures varied widely, with only tick checks regularly applied, while protective clothing and repellents were often ignored or not applied consistently ([11, 13, 14]). TBE vaccination uptake remains low across Europe, including Switzerland ([15]), with vaccine skepticism, lack of information, and low risk perception identified as the main barriers ([16]). KAP studies in non-endemic regions have shown heterogeneous knowledge of arboviral diseases and only moderate adoption of protective measures, even in areas where *Ae. albopictus* is established ([17]). Besides mosquito biting frequency, concern about contracting diseases and socioeconomic status are associated with protective behaviour ([17, 18]). No study has examined KAP regarding both tick-borne and mosquito-borne VBDs jointly in the Swiss general population, and no data are available on awareness or preparedness in relation to the evolving arboviral risk in Switzerland.

To address these gaps, we conducted two complementary surveys within the Swiss National Centre for Climate Services (NCCS)-Impacts programme. The first one was a population-based survey embedded in the BEready cohort in the canton of Bern, designed to assess KAP regarding VBDs among adults. A second survey collected information on institutional engagement, existing measures, perceived gaps, and coordination around VBDs among local authorities across the human health, animal health, and environmental sectors in Switzerland and Liechtenstein. This stakeholder survey is used here as additional material to help interpret the population-level findings. Individual behaviours remain central to the prevention of both tick- and mosquito-borne infections, and public acceptance of surveillance and control measures can influence the feasibility of broader public health action. Understanding where knowledge levels are adequate and where gaps persist is therefore relevant for both risk communication and climate-sensitive adaptation planning. The main objective of this study was to describe KAP regarding VBDs among adults in the canton of Bern in 2025. We further aimed to characterize the latent structure of VBD knowledge using item response theory, examine sociodemographic and exposure-related correlates of latent knowledge, identify KAP profiles through exploratory clustering, and interpret these population findings in light of complementary evidence on institutional engagement from the stakeholder survey.

## 2 Methods

### 2.1 General population KAP data

We conducted a cross-sectional, web-based survey of adults (*≥* 18 years) residing in Switzerland in 2025. The survey was implemented within the BEready platform (University of Bern/Multidisciplinary Center for Infectious Diseases), which is registered on ClinicalTrials.gov (ID: NCT06739499) and operates as a general population-based prospective cohort in the canton of Bern under a One-Health framework. The cohort has enrolled 1,569 households, including adults, children and pets, recruited by random selection from the cantonal residents’ register and supplemented by volunteer households. Between April and September 2023, a pilot study with 108 households was conducted [19]. Enrolment for the main study started in April 2024 and was completed in January 2026. A module on VBDs was added to the 1-year follow-up questionnaire of pilot-study participants, and to the baseline questionnaire of main-study participants. Data from all participants completing the VBD module up to May 2025 were included in the present analysis.

Eligible participants were adults (*≥* 18 years) who provided informed consent, resided on average more than three days per week in a private household in the canton of Bern, had access to the internet, held a personal email address, and had sufficient proficiency in German, French, or English to complete the survey. Individuals were excluded if they had a planned move outside the canton of Bern during the study period. At inclusion, participants undergo a brief clinical examination and provide a blood sample, then complete an online questionnaire covering health status, infection susceptibility, and KAP concerning VBDs. The study was approved by the responsible ethics committee (Business Administration System for Ethics Committees (BASEC) numbers 2023-00333 and 2023-02290) and is conducted in accordance with Swiss legislation and applicable international guidelines.

The KAP questionnaire follows a structured, multi-stage approach to assess knowledge about VBDs and their transmission, incorporating both familiarity and understanding. In the first stage, respondents were presented with three items to determine whether they had previously encountered the relevant concepts or understood the underlying concepts as follows. 1) An introductory text provided respondents with a definition of VBDs and the fact that tick-borne encephalitis is a disease transmitted by ticks. Respondents were then asked to classify tick-borne encephalitis as vector-borne or not, serving as an initial measure of their conceptual understanding. 2) A disease familiarity component, capturing whether respondents reported prior exposure to the names of particular diseases. 3) A vector familiarity component, capturing whether they reported prior exposure to the names of specified organisms. In the second stage, participants completed classification exercises in which they were required to identify, from structured lists, which diseases they believed were vector-borne and which organisms functioned as vectors.

Establishing the first stage baseline was essential to distinguish between lack of knowledge and misunderstanding in subsequent tasks. To evaluate the robustness of conceptual understanding and to detect systematic misconceptions, distractor items were intentionally included: measles and influenza in the disease set, and wasps and bed bugs in the vector set. These items, while familiar to most respondents, do not fall within the epidemiological definition of VBDs or recognized disease vectors, and thus provided a validity check. Results from the first stage as well as logical contradictions in the second stage were used to filter out unreliable answers.

The questionnaire also assessed self-reported exposure, preventive practices, perceived disease relevance, and beliefs about public control measures. Exposure was measured by reported annual tick-bite frequency and summer mosquito-bite frequency. Preventive practices were assessed separately for ticks and mosquitoes, including personal protective behaviours, environmental or household measures, absence of active measures, and open-text responses for other practices. Attitudes were assessed through perceived effectiveness of tick- and mosquito-control measures, perceived current relevance of selected diseases as health problems in Switzerland, and perceived need for vector-control measures in Switzerland.

To enhance the reliability of downstream KAP analyses, the knowledge section incorporated predefined checks for conceptual understanding and internal consistency. These checks included distractor items assessing whether respondents could distinguish vector-borne transmission from familiar but non-vector-borne diseases or non-vector organisms. Influenza and measles were included as non-vector-borne disease distractors, and wasps and bed bugs as non-vector organism distractors. Before analysis, we applied two predefined validity criteria to identify responses that were sufficiently interpretable for KAP analysis. Respondents were retained in the primary analysis sample if they classified tick-borne encephalitis as a VBD after the introductory definition and if they showed no logical contradiction between vector-recognition and disease/vector association items.

The online questionnaire was implemented using Research Electronic Data Capture (REDCap) tools hosted at the University of Bern [20, 21]. The full questionnaire is provided in Supplementary Appendix 1. All subsequent analysis and description of the data was performed in R (version 4.2.0). Descriptive summaries of participant characteristics and questionnaire responses were based on observed data in the primary analysis sample; missing questionnaire responses were retained and displayed explicitly where relevant. For model-based analyses and clustering, missing values in variables required for the IRT model, regression models, and PAM clustering were imputed after application of the validity criteria. Imputation was performed using iterative chained imputation with gradient-boosted decision trees implemented with the R package xgboost [22].

### 2.2 Latent knowledge modelling using item response theory

To obtain a latent summary of participants’ overall knowledge of VBDs, we fitted an item response theory (IRT) model to the knowledge items. The objective of this analysis was to derive a continuous respondent-level knowledge score covering all questions, that accounts for differences between items in difficulty and informativeness. Each knowledge item was coded as correct or incorrect. “Do not know” responses were coded as incorrect, because they indicate absence of the targeted knowledge.

We used a two-parameter logistic IRT model [23]. For participant *i* and item *j*, the probability of a correct response was modelled as

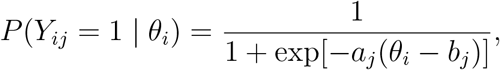

where *θ_i_* denotes the latent overall knowledge level of participant *i*, *a_j_* is the discrimination parameter of item *j*, and *b_j_* is the difficulty parameter of item *j*. The discrimination parameter quantifies how strongly an item differentiates participants along the latent knowledge continuum, whereas the difficulty parameter indicates the level of latent knowledge at which the probability of a correct response reaches 0.5. Parameter estimation was performed using quadrature-based expectation–maximization (EM) algorithm using the R package mirt ([24]).

The initial candidate set of knowledge items comprised disease-classification items, disease/vector association items, and vector-recognition items. During model development, the vector-recognition items, which asked participants whether specific organisms can transmit diseases, were excluded from the knowledge score. For non-vector distractor organisms, response patterns were nearly invariant and discrimination estimates were close to zero. Conversely, true vector organisms, particularly ticks and mosquitoes, were correctly identified by almost all participants, resulting in quasi-complete separation and unstable item-parameter estimation. Because these items impaired model convergence and latent score estimation, the final IRT score was restricted to 7 disease-classification and 8 disease/vector association items.

Participant-level latent knowledge scores were extracted from the fitted model and standardized for subsequent analyses. Higher values indicate greater estimated knowledge of VBDs. These scores were used both descriptively and as outcomes in regression models assessing associations between knowledge and participant-level variables.

### 2.3 Determinants of latent knowledge

We examined factors associated with participants’ latent overall knowledge of VBDs using regression models with the standardized IRT-derived knowledge score as the outcome. Because the score represents a continuous estimate of each participant’s position on the latent knowledge continuum, we used ordinary least squares (OLS) linear regression models to quantify associations between participant characteristics, exposure history, geographical context, travel history, and knowledge.

The model included sociodemographic variables, self-reported vector exposure, municipality topography, and travel-related variables. Age was modelled as a continuous predictor using a linear term, expressed per 10-year increase after centering at the sample mean, and a quadratic term based on the same centered 10-year scale to allow for non-linear associations with knowledge. Sex, education level, residence history, tick-bite frequency, mosquito-bite frequency, municipality topography, and visited continents were included as categorical predictors. Educational attainment was collapsed from the original questionnaire categories into four groups adapted to the Swiss education system: compulsory education, upper secondary education, tertiary professional education, and tertiary university education. Self-reported mosquito-bite frequency during summer was recoded so that the categories "I don’t know", "Never", and "Less than yearly" were combined into a single reference category, labelled "Unknown/exceptional".

### 2.4 Clustering procedure

We performed an exploratory clustering analysis to identify profiles of participants with similar KAP regarding VBDs. The analysis was based on the completed questionnaire dataset. Only variables directly measuring knowledge, perceived disease relevance, perceived effectiveness of vector-control measures, and self-reported preventive practices were used as active clustering variables. Sociodemographic variables, municipality topography, self-reported tick and mosquito exposure, and travel history were excluded from the clustering algorithm and used only to describe the resulting clusters.

Because the questionnaire contained unequal numbers of items across domains, active variables were grouped into conceptual blocks before computing participant dissimilarities. The blocks were: knowledge, tick-related preventive practices, mosquito-related preventive practices, perceived relevance of tick-borne diseases, perceived relevance of mosquito-borne diseases, perceived effectiveness of tick-control measures, and perceived effectiveness of mosquito-control measures. Pairwise dissimilarities were calculated using a block-weighted Gower distance. Within each block, item-level Gower dissimilarities were averaged across the variables in that block. The final dissimilarity between two participants was then defined as the average of the block-specific dissimilarities. This weighting scheme allowed binary and categorical questionnaire variables to be combined while preventing domains with more items from dominating the clustering solution.

Clustering was performed using partitioning around medoids (PAM) applied to the block-weighted Gower dissimilarity matrix. The number of clusters was fixed at *k* = 6 after considering internal validity indices, cluster stability, minimum cluster size, and substantive interpretability. In comparative diagnostics, PAM solutions based on Gower dissimilarities performed better than the alternative approaches considered, including k-means and Ward hierarchical clustering, and the block-weighted Gower distance performed better than the unweighted Gower distance. The final clusters were characterized using domain-level mean scores for the active variables and, post hoc, by sociodemographic characteristics, municipality topography, vector exposure, and travel history.

### 2.5 Stakeholder survey

To assess the engagement of cantonal and Liechtenstein authorities with regard to VBDs, a cross-sectional survey was conducted among stakeholders in departments responsible for health, veterinary services, and environment. In order to facilitate open communication and increase willingness to participate, the responses were not attributed to specific locations or departments outside of the general area of human health, animal health and environment. Consequently, our analysis is limited in that it does not identify which canton or department provided specific information, nor does it allow for comparison between cantonal approaches. The stakeholder survey was administered as an online questionnaire through REDCap, hosted by Unisanté. The questionnaire was distributed by email to targeted respondents within relevant services, based on their role and responsibilities in managing public health or environmental risks. Participation was voluntary.

The online survey was open from April 25 to June 6, 2025. A reminder was sent on May 16. The questionnaire included structured multiple-choice and checkbox items, as well as a small number of open-ended questions, covering topics such as awareness of VBDs, existing preventive measures, interdepartmental collaboration, and perceived needs or challenges. Respondents were assured that they would not be identified.

The full questionnaire is available online in supplementary appendix 2. Data were anonymised, exported securely from REDCap and described using R (version 4.4.2). Open-text answers in multiple languages were summarized using a locally run large language model (qwen3.6:35b, june 2026). More details are available in the supplementary file.

## 3 Results

### 3.1 Participant characteristics

Among 1,847 respondents from the BEready cohort, 1,415 correctly classified tick-borne encephalitis as a VBD in the initial conceptual validity item and 1,604 provided internally consistent answers across the vector-recognition and disease/vector matching items. The primary analysis sample comprised 1,337 respondents who both passed the conceptual screen and provided logically consistent responses.

The mean age of participants in the primary analysis sample was 53.4 years (SD 15.5). The sample included 765 women (57.2%) and 572 men (42.8%). Educational attainment was high: among 1,294 participants with non-missing education, 651 (50.3%) had tertiary university education and 280 (21.6%) had tertiary professional education, whereas 343 (26.5%) had upper-secondary education and 20 (1.5%) had compulsory education only. Most participants had lived in Switzerland since birth (970/1,334, 72.7%), while 278 (20.8%) were born abroad and 86 (6.4%) were Swiss-born but had lived abroad. Most lived in urban municipalities (873/1,330, 65.6%), followed by intermediate (300/1,330, 22.6%) and rural municipalities (157/1,330, 11.8%).

Self-reported exposure differed substantially between ticks and mosquitoes. Among 1,332 participants with non-missing tick-bite frequency, 784 (58.9%) reported no tick bites in a usual year and 471 (35.4%) reported 1–3 tick bites; 77 (5.8%) reported four or more tick bites per year. Mosquito bites were more frequent: among 1,310 respondents with non-missing mosquito-bite frequency, 1,085 (82.8%) reported being bitten at least monthly during summer, and 692 (52.8%) at least weekly.

Across the 79 variables selected for imputation, 760 of 105,623 values were imputed (0.7%). These missing values occurred in 330 participants, corresponding to 24.7% of the primary analysis sample. Missingness was concentrated in disease/vector matching items, which accounted for 667 of 760 imputed values (87.8%). The largest numbers of imputed values were observed for chikungunya/mosquitoes (154/1,337, 11.5%), measles/no vector (138/1,337, 10.3%), influenza/no vector (135/1,337, 10.1%), West Nile fever/mosquitoes (117/1,337, 8.8%), and Zika/mosquitoes (77/1,337, 5.8%). Additional imputed values concerned education (43/1,337, 3.2%), mosquito-bite frequency (27/1,337, 2.0%), dengue/mosquitoes (22/1,337, 1.6%), Lyme disease/ticks (17/1,337, 1.3%), tick-borne encephalitis/ticks and municipality topography (7/1,337 each, 0.5%), tick-bite frequency (5/1,337, 0.4%), perceived need for vector-control measures in the raw and recoded variables (4/1,337 each, 0.3%), and Swiss residence history (3/1,337, 0.2%). After imputation, the variables required for the IRT, regression, and PAM clustering analyses were complete.

**Table 1:** Characteristics of participants included in the primary analysis sample.

| Characteristic | Overall |
| --- | --- |
| **Age, years** |  |
| Mean (SD), n = 1,337 | 53.4 (15.5) |
| **Sex, n = 1,337** |  |
| Male | 572 (42.8%) |
| Female | 765 (57.2%) |
| **Education, n = 1,294** |  |
| Compulsory | 20 (1.5%) |
| Upper secondary | 343 (26.5%) |
| Tertiary professional | 280 (21.6%) |
| Tertiary university | 651 (50.3%) |
| **Residence history, n = 1,334** |  |
| Since birth | 970 (72.7%) |
| Swiss-born, lived abroad | 86 (6.4%) |
| Born abroad | 278 (20.8%) |
| **Municipality topography, n = 1,330** |  |
| Urban | 873 (65.6%) |
| Intermediate | 300 (22.6%) |
| Rural | 157 (11.8%) |
| **Tick-bite frequency, n = 1,332** |  |
| 0 | 784 (58.9%) |
| 1–3 | 471 (35.4%) |
| 4–6 | 38 (2.9%) |
| 7–9 | 15 (1.1%) |
| ≥10 | 24 (1.8%) |
| **Mosquito-bite frequency, n = 1,310** |  |
| Unknown/exceptional | 103 (7.9%) |
| At least yearly | 122 (9.3%) |
| At least monthly | 393 (30.0%) |
| At least weekly | 573 (43.7%) |
| Every day | 119 (9.1%) |

Values are n (%) unless otherwise stated. Percentages are calculated among participants with non-missing values for each variable.

### 3.2 Knowledge, attitudes and practices

Knowledge was strongest for the recognition of major vectors and for tick-related disease– vector associations (Figure 1). Ticks were identified as vectors by 1,336 participants (99.9%) and mosquitoes by 1,319 (98.7%). Tick-borne encephalitis was matched to ticks by 1,310 participants (98.0%), and Lyme disease was matched to ticks by 1,159 (86.7%). Correct disease classification was high for dengue (1,164, 87.1%) and Lyme disease (1,115, 83.4%), intermediate for Zika (939, 70.2%), and lower for West Nile fever (577, 43.2%) and chikungunya (475, 35.5%). A similar pattern was observed for disease–vector matching: dengue was matched to mosquitoes by 1,150 participants (86.0%), Zika by 906 (67.8%), West Nile fever by 731 (54.7%), and chikungunya by 529 (39.6%).

**Figure 1:**
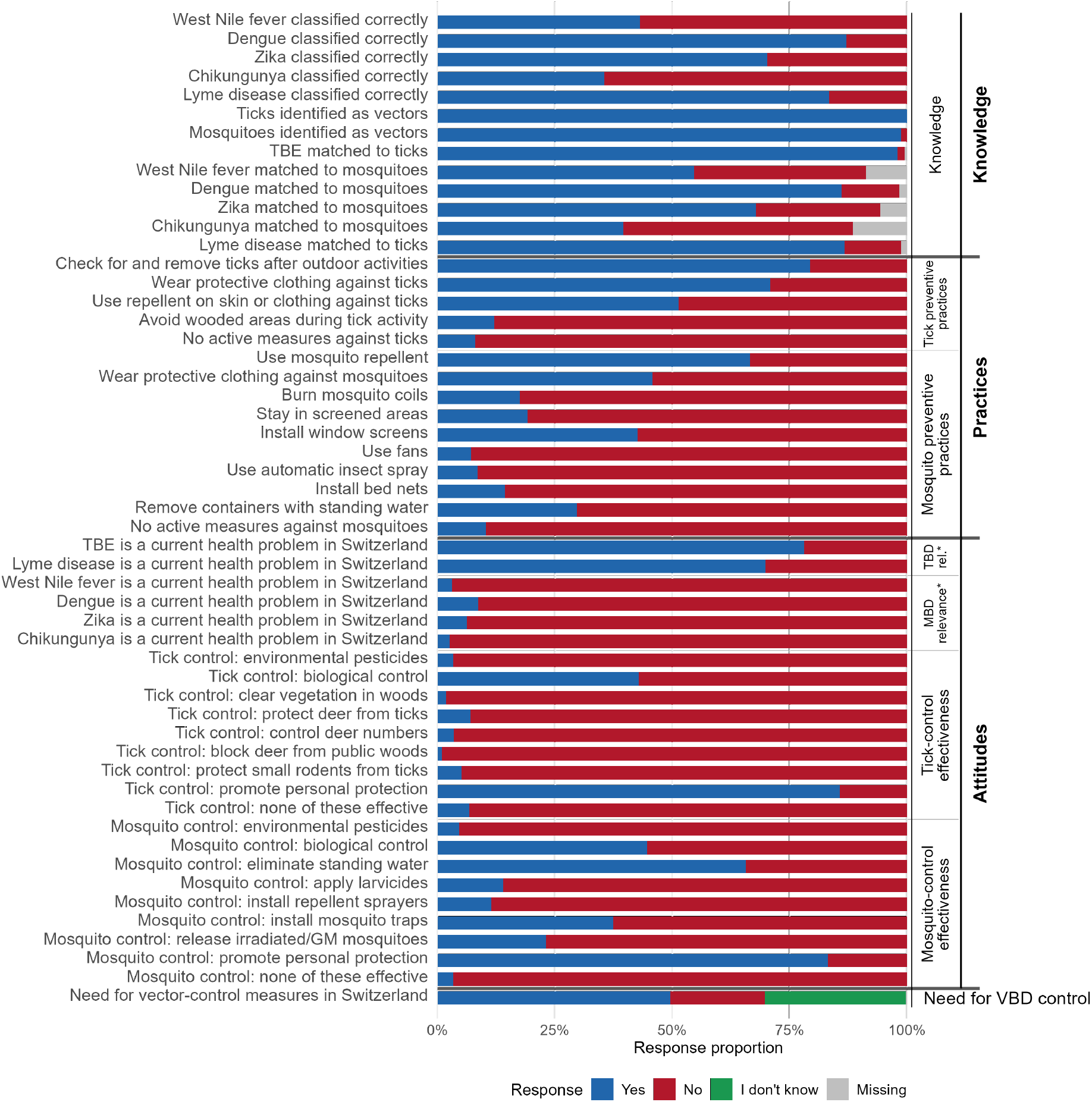
Observed questionnaire response proportions in the primary analysis sample across knowledge, attitudes, practices, and perceived need for vector control. For knowledge items, “Yes” denotes a correct answer or correct association. For practice and attitude items, “Yes” denotes endorsement or selection of the item. Missing responses are displayed as a separate category.

Preventive practices were more frequently reported for ticks than for mosquitoes. For ticks, 1,060 participants (79.3%) reported checking for and removing ticks after outdoor activities, 947 (70.8%) reported wearing protective clothing, and 686 (51.3%) reported using repellent on skin or clothing. Avoidance of wooded areas during periods of high tick activity was less frequent (161/1,337, 12.0%), and 107 participants (8.0%) reported taking no active tick-prevention measures. For mosquitoes, the most common preventive measures were repellent use (889, 66.5%), protective clothing (612, 45.8%), and window screens (569, 42.6%). Removal of containers with standing water was reported by 396 participants (29.6%), and 138 (10.3%) reported taking no active measures against mosquitoes.

Perceived disease relevance was markedly higher for tick-borne than mosquito-borne diseases. Tick-borne encephalitis was considered a current health problem in Switzerland by 1,044 participants (78.1%), and Lyme disease by 933 (69.8%). By contrast, perceived relevance was low for dengue (115, 8.6%), Zika (83, 6.2%), West Nile fever (41, 3.1%), and chikungunya (34, 2.5%). Perceived effectiveness of control measures was concentrated on personal protection and selected simple environmental measures. Promoting personal protection was endorsed by 1,146 participants (85.7%) for tick control and 1,112 (83.2%) for mosquito control. Biological control was endorsed by 572 participants (42.8%) for ticks and 597 (44.7%) for mosquitoes. Elimination of standing water was endorsed by 878 participants (65.7%) for mosquito control, whereas more technical or ecological measures were less frequently selected. Overall, 662 participants (49.7%) considered vector-control measures necessary in Switzerland, 270 (20.3%) did not, and 401 (30.1%) answered “don’t know”.

### 3.3 Patterns in knowledge

The two-parameter logistic IRT model was fitted to 15 disease-classification and disease– vector association items after imputation of missing analytical variables (Figure 2A).

**Figure 2:**
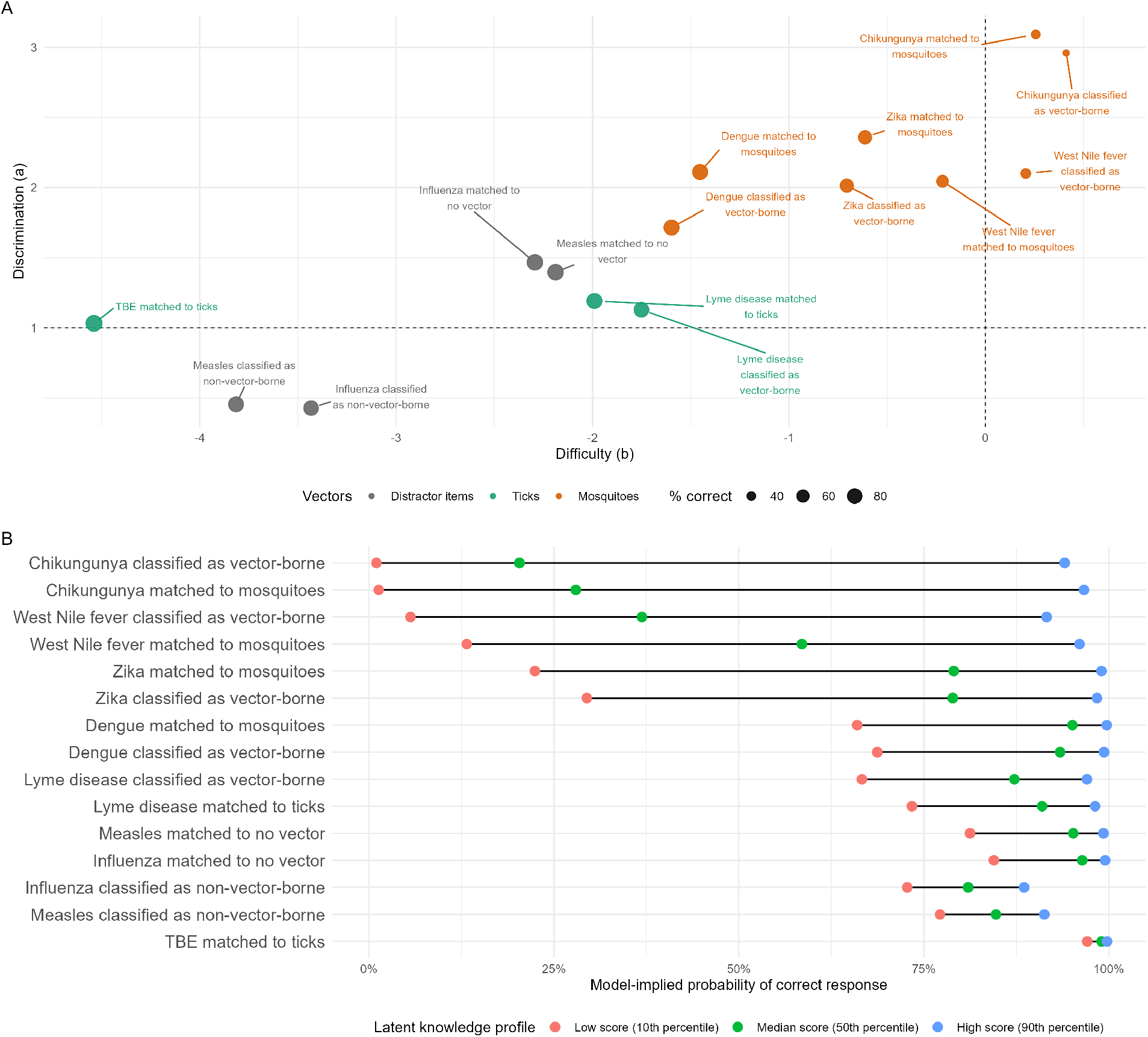
Item response theory model for VBD knowledge. Panel A shows item discrimination (parameter *a*, higher value means higher discrimination) and difficulty (parameter *b*, higher value means higher difficulty) from the two-parameter logistic model, with point size proportional to the proportion of correct responses and colour indicating item group. Panel B shows model-implied probabilities of correct response for low, median, and high latent knowledge profiles.

The item-level parameter estimates showed a clear distinction between easier tick-related or distractor items and more difficult mosquito-borne disease items. The easiest item was matching tick-borne encephalitis to ticks (difficulty *b* = *−*4.54; 98.5% correct). Other low-difficulty items included classification of measles as non-vector-borne (*b* = *−*3.81; 84.1% correct), classification of influenza as non-vector-borne (*b* = *−*3.43; 80.5% correct), matching influenza to no vector (*b* = *−*2.29; 93.0% correct), and matching measles to no vector (*b* = *−*2.19; 91.5% correct). Lyme disease items were also relatively easy: matching Lyme disease to ticks had difficulty *b* = *−*1.99 and classification of Lyme disease as vector-borne had difficulty *b* = *−*1.75.

The most difficult items were mosquito-borne disease items. Chikungunya classification as vector-borne had the highest difficulty (*b* = 0.41; 35.5% correct), followed by matching chikungunya to mosquitoes (*b* = 0.26; 40.4% correct) and classifying West Nile fever as vector-borne (*b* = 0.21; 43.2% correct). These items also had high discrimination: matching chikungunya to mosquitoes (*a* = 3.09), classifying chikungunya as vector-borne (*a* = 2.96), matching Zika to mosquitoes (*a* = 2.36), and classifying West Nile fever as vector-borne (*a* = 2.10) were among the most informative items for separating respondents along the latent knowledge continuum. Overall, the IRT model indicated that knowledge gaps were concentrated in less familiar mosquito-borne diseases, whereas tick-related items and non-vector distractors were generally easier.

### 3.4 Determinants of latent knowledge score

In the multivariable linear model, the outcome was the standardized IRT-derived latent knowledge score (Figure 3). The model included 1,337 participants and explained a modest proportion of between-participant variation in knowledge (R^2^=0.099; adjusted R^2^=0.082).

**Figure 3:**
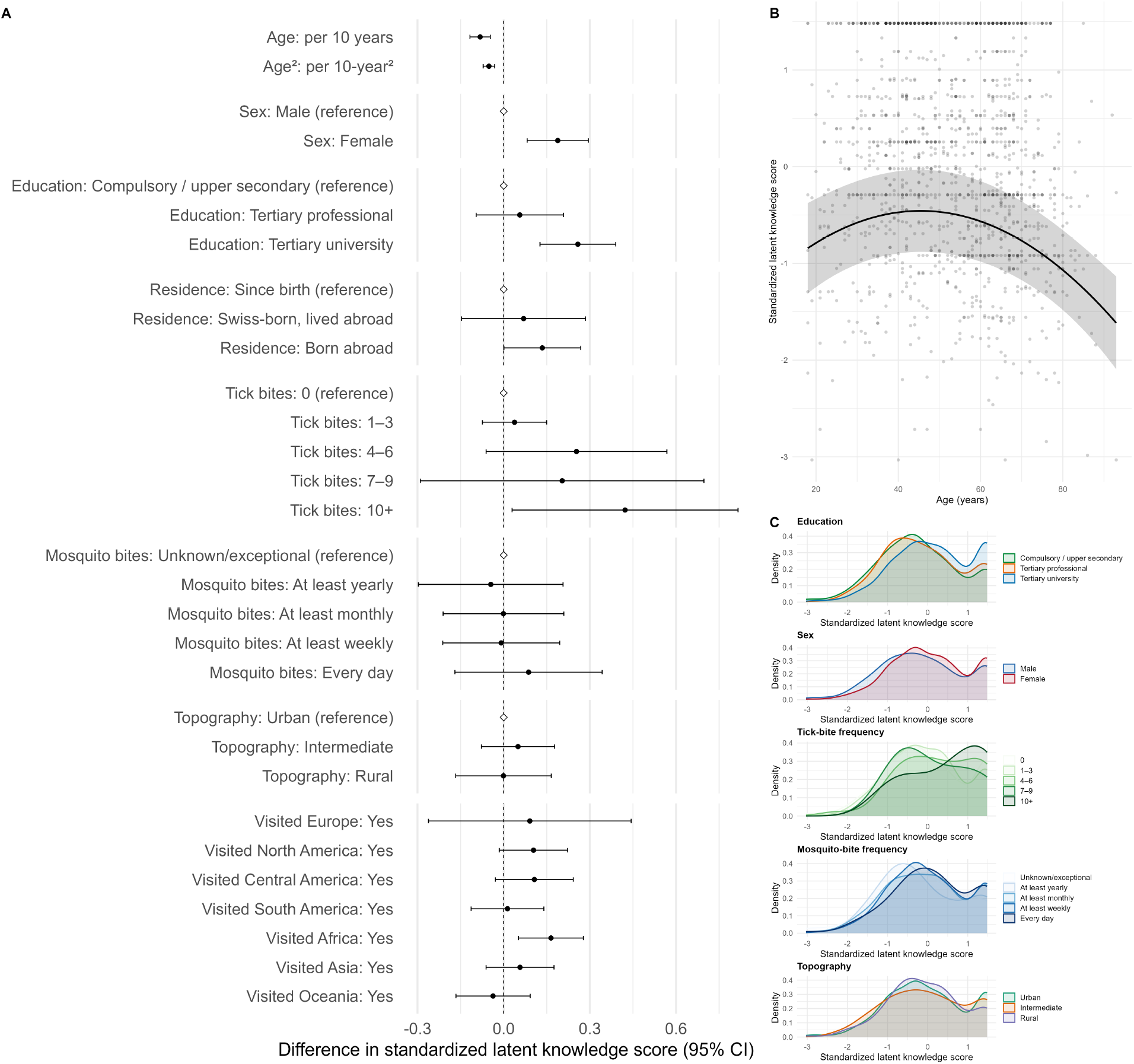
Determinants of the standardized IRT-derived latent knowledge score. Panel A shows adjusted regression coefficients with 95% confidence intervals; diamonds indicate reference categories at zero. Age is displayed as a linear term per 10 years and a quadratic term per squared 10-year unit. Panel B shows the adjusted age curve with empirical participant-level data. Panel C shows unadjusted density distributions of the latent knowledge score by selected categorical variables.

Age showed a non-linear association with latent knowledge. With age centred at the sample mean and expressed per 10 years, the linear age coefficient was *β* = *−*0.082 standard deviations per 10 years (95% CI -0.118 to -0.047), and the quadratic term was *β* = *−*0.051 per squared 10-year unit (95% CI -0.071 to -0.031). The fitted curve suggested highest adjusted knowledge in mid-adulthood, followed by lower predicted knowledge at older ages.

Female participants had higher latent knowledge than male participants (*β* = 0.188, 95% CI 0.081 to 0.294). Compared with participants who had lived in Switzerland since birth, those born abroad had slightly higher latent knowledge (*β* = 0.136, 95% CI 0.002 to 0.270), whereas the estimate for Swiss-born participants who had lived abroad was compatible with no association (*β* = 0.069, 95% CI -0.147 to 0.285).

Educational attainment was associated with latent knowledge. Because the compulsory education group was small, compulsory and upper-secondary education were combined as the reference category. Compared with participants with compulsory or upper-secondary education, those with tertiary university education had higher latent knowledge (*β*=0.258, 95% CI 0.126 to 0.390), whereas the estimate for tertiary professional education was smaller and compatible with no association (*β*=0.056, 95% CI -0.096 to 0.208).

Tick exposure showed a positive association only at the highest exposure level. Compared with participants reporting no tick bites, those reporting 10 or more tick bites per year had higher latent knowledge (*β* = 0.424, 95% CI 0.030 to 0.818). Estimates for 1–3, 4–6, and 7–9 tick bites per year were positive but compatible with no association. Mosquito-bite frequency was not significantly associated with knowledge after adjustment. Municipality topography was also not significantly associated with knowledge. Among travel variables, having visited Africa was associated with higher latent knowledge (*β* = 0.165, 95% CI 0.051 to 0.278), whereas estimates for other continents were smaller and compatible with no significant association.

### 3.5 Knowledge–attitude–practice clusters

The block-weighted Gower/PAM analysis identified six clusters of participants based on knowledge, tick and mosquito preventive practices, perceived disease relevance, and perceived effectiveness of control measures (Figure 4). Cluster sizes ranged from 170 to 286 participants. Cluster 1 included 170 participants (12.7%), Cluster 2 included 217 (16.2%), Cluster 3 included 235 (17.6%), Cluster 4 included 286 (21.4%), Cluster 5 included 215 (16.1%), and Cluster 6 included 214 (16.0%).

**Figure 4:**
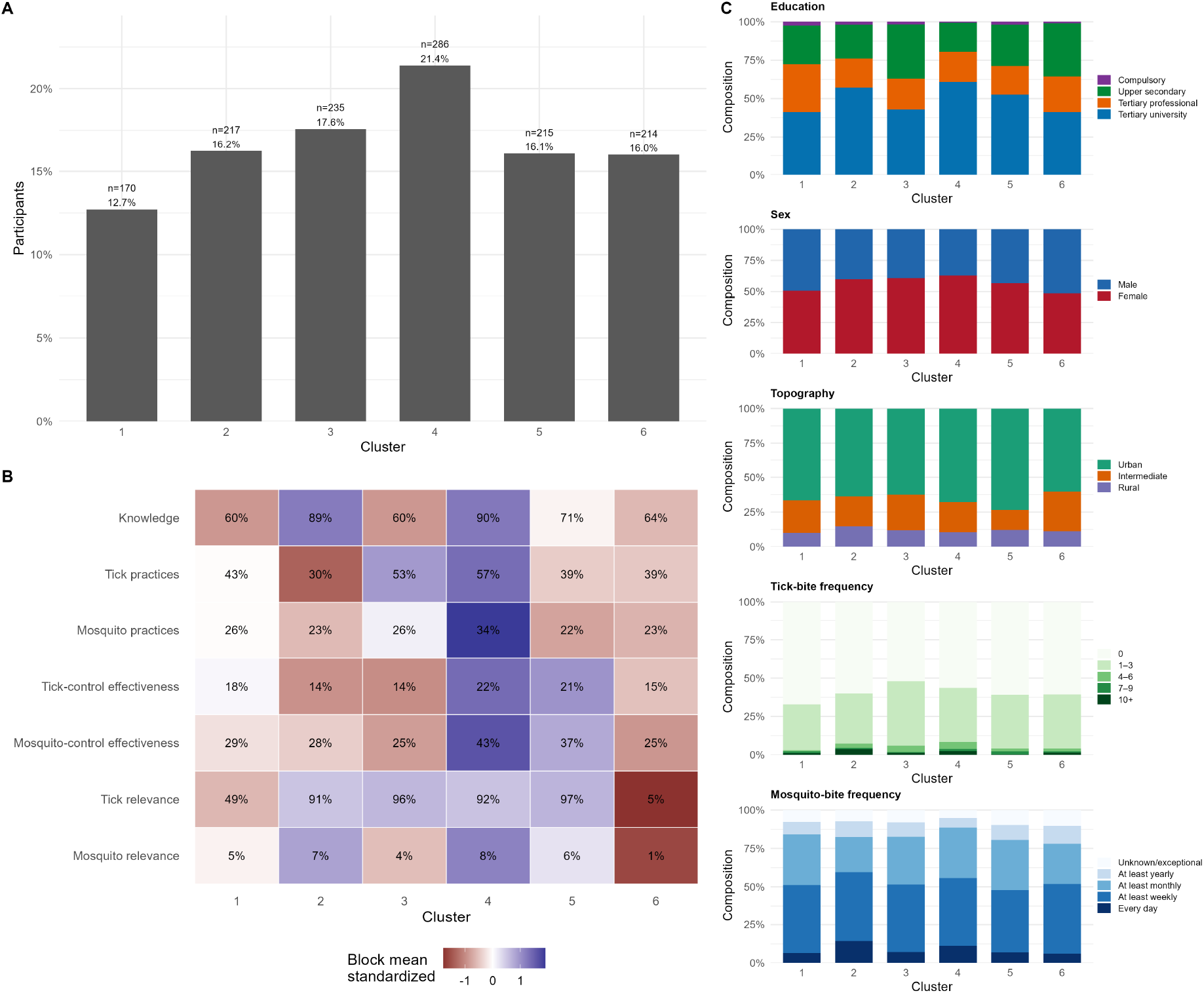
Knowledge–attitude–practice clusters from partitioning around medoids applied to the block-weighted Gower distance. Panel A shows cluster size. Panel B shows cluster-level mean block scores; numbers inside tiles are raw mean block scores, while colors represent within-block standardized relative values across clusters. Panel C shows cluster composition by education, sex, municipality topography, tick-bite frequency, and mosquito-bite frequency.

The clusters differed primarily in their KAP profiles. Cluster 4 showed the most consistently high engagement profile, with high knowledge (mean block score 90%), high tick-related prevention practices (57%), the highest mosquito-related prevention practices (34%), high tick relevance (92%), the highest mosquito relevance (8%), and the highest perceived effectiveness of both tick-control (22%) and mosquito-control measures (43%). Cluster 2 also showed high knowledge (89%) and high perceived tick relevance (91%), but lower reported tick practices (30%) and mosquito practices (23%). Cluster 3 combined lower knowledge (60%) with high perceived tick relevance (96%) and relatively frequent tick practices (53%), but lower perceived effectiveness of both tick-control (14%) and mosquito-control measures (25%). Cluster 5 had intermediate knowledge (71%), the highest perceived tick relevance (97%), and relatively high perceived effectiveness of mosquito-control measures (37%), but lower mosquito practices (22%). Cluster 1 had lower knowledge (60%), moderate tick relevance (49%), and intermediate prevention scores. Cluster 6 was characterized by low perceived relevance, particularly for tick-borne diseases (5%) and mosquito-borne diseases (1%), together with lower knowledge (64%), lower mosquito practices (23%), and lower perceived effectiveness of mosquito-control measures (25%).

The sociodemographic composition of clusters was broadly similar, although some differences were apparent. Tertiary university education was most frequent in Cluster 4 (174/286, 60.8%) and Cluster 2 (124/217, 57.1%), and least frequent in Cluster 6 (88/214, 41.1%) and Cluster 1 (70/170, 41.2%). Women were the majority in Clusters 2–5, with the highest proportion in Cluster 4 (180/286, 62.9%), whereas Cluster 6 had a slight male majority (110/214, 51.4%). Urban residence predominated in all clusters, ranging from 129/214 (60.3%) in Cluster 6 to 158/215 (73.5%) in Cluster 5. Exposure distributions were also broadly similar across clusters: in every cluster, the most common tick-bite category was no tick bites, and the most common mosquito-bite category was at least weekly bites.

### 3.6 Complementary stakeholder findings

#### 3.6.1 Participants

We conducted a nationwide survey across all cantons, contacting 153 email addresses from the health, environment and veterinary departments. Some recipients responded indicating their department was not responsible for the survey topics, and others redirected us to appropriate departments. Ultimately, we received 55 complete responses, covering all 26 Swiss cantons and Liechtenstein. The number of answers per canton varied between 1 and 3. Participants primarily belonged to human health cantonal departments (n= 22), followed by the animal health departments (n= 17) and the environment departments (n= 16).

#### 3.6.2 Involvement and institutional capacity

Most participants (77%) reported that their authority is currently involved in the implementation of measures or activities related to diseases transmitted by ticks and mosquitoes, such as public relations, monitoring, or control (Supplementary Table S2.1). This was especially true in animal health departments (94%). Regarding the existence of cantonal strategies or action plans, 49% of respondents were aware of a plan for mosquito-borne diseases and 11% for tick-borne diseases, while 33% reported that no strategy exists in their canton. Many authorities reported limited dedicated resources: 49% indicated that their authority had no human resources specifically allocated to activities related to tick- or mosquito-borne diseases, and 55% reported that no dedicated budget was available for such activities. Among 12 of 55 respondents who indicated no current involvement, the most frequently cited reasons included a lack of financial or human resources (6 out of 12) and a perception that such diseases are outside the authority’s responsibilities (5 out of 12).

The stakeholder survey revealed a marked asymmetry between institutional responses to mosquito-borne and tick-borne diseases. Although 77% of respondents reported that their authority was involved in VBD activities, formal strategies were far more often reported for mosquito-borne diseases (49%) than for tick-borne diseases (11%). Concrete mosquito-related activities were also broader and more operational: authorities reported active and passive monitoring, verification of new sightings, public communication, professional awareness, and, in some cases, mosquito control on public or private land. By contrast, tick-related engagement was concentrated mainly in public information and professional awareness, with very limited monitoring or operational control.

A second notable finding is that this broader mosquito-related engagement is occurring despite limited institutional capacity. Roughly half of respondents reported no dedicated human resources (49%) and no dedicated budget (55%) for VBD activities. Among authorities not currently involved, the main reasons were lack of resources and uncertainty about whether the issue fell within their mandate. Respondents also identified important knowledge gaps, especially for mosquito-borne diseases, in vector and disease management, prevention, detection, and long-term control. Uncertainty around responsibilities, the use of adulticides, and coordination across actors emerged repeatedly, indicating that operational ambition is outpacing institutional capacity and support.

Finally, the survey highlighted the importance of cross-sectoral coordination. Collaboration with researchers and specialists was much more common for mosquito-borne diseases (60%) than for tick-borne diseases (20%), and environmental services were particularly active in mosquito surveillance and control, whereas human health services were more prominent in tick-related communication. At the same time, many respondents reported limited awareness of whether VBDs were integrated into national or cantonal climate adaptation strategies.

## 4 Discussion

### 4.1 Findings

In this population-based survey of adults in the canton of Bern, KAP regarding VBDs showed a clear asymmetry between tick-borne and mosquito-borne risks. Public knowledge was stronger for tick-borne diseases, especially TBE and Lyme disease, which were widely recognized and commonly perceived as relevant to Switzerland. By contrast, knowledge of mosquito-borne diseases was less complete, particularly for West Nile fever and chikungunya, even though mosquito exposure was frequent in daily life.

Protective behaviours were common. Participants reported high uptake of several recommended preventive measures, particularly for ticks. For mosquitoes, respondents also commonly reported using repellents and window screens, although removal of standing water was less frequent. Support for control measures centred on personally actionable and relatively simple interventions, whereas support for more complex or less familiar strategies was limited.

The exploratory multivariable analysis of the IRT-derived latent knowledge score identified several participant-level correlates of knowledge. Tertiary university education was associated with higher latent knowledge compared with compulsory or upper-secondary education, whereas the estimate for tertiary professional education was smaller and compatible with no clear association. Female sex, being born abroad, reporting ten or more tick bites per year, and having visited Africa were also associated with higher latent knowledge. Age showed a non-linear association, with lower adjusted knowledge at older ages.

The exploratory clustering analysis additionally showed that KAP profiles were heterogeneous. Some clusters combined high knowledge with high perceived tick-borne disease relevance but comparatively lower preventive practices, whereas others showed lower knowledge but relatively high tick-related prevention or stronger endorsement of mosquito-control measures. Sociodemographic differences between clusters were present but moderate, suggesting that the clusters mainly captured differences in engagement, perceived relevance, and control attitudes rather than sharply separated population subgroups.

The complementary stakeholder survey suggested a contrasting institutional pattern. Mosquito-borne threats appeared to be addressed as a more complex, emerging, and multisectoral preparedness issue, with broader engagement in surveillance, monitoring, and operational control, whereas tick-borne diseases appeared to be managed mainly through public information, professional awareness, and vaccination-oriented prevention. However, preparedness remained constrained by fragmented responsibilities, uneven coordination, and limited dedicated human and financial resources.

### 4.2 Interpretation

These findings suggest that the public in Bern is relatively well prepared for familiar tickborne risks but less prepared, from a knowledge perspective, for mosquito-borne threats that may become increasingly relevant under changing climatic conditions. The fact that mosquito bites were common while mosquito-borne disease knowledge remained comparatively incomplete is epidemiologically important. Familiarity with exposure does not appear to translate into familiarity with disease risk. Rather, the public appears more familiar with established tick-borne risks than with mosquito-borne threats that remain less visible in the Swiss context but may become increasingly relevant in the future.

The complementary stakeholder findings sharpen this interpretation. Cantonal and Liechtenstein authorities reported broader and more diversified engagement for mosquitoborne diseases than for tick-borne diseases, especially in surveillance and control-related activities. This may reflect the novelty, operational complexity, and cross-sectoral nature of mosquito-related threats, including invasive species monitoring and local control activities. Yet public knowledge remains more established for tick-borne diseases. The resulting contrast suggests a preparedness gap: institutional concern and activity are relatively developed for mosquito-related threats, but population-level literacy lags behind.

This mismatch has practical implications. Institutional readiness alone may not be sufficient if household- and community-level measures depend on public understanding of why mosquito-borne diseases matter in the Swiss context. Conversely, strong public familiarity with tick-borne risks may partly reflect long-standing communication, visible public health messaging, and a clearer domestic disease narrative.

The cluster analysis further suggests that public-health communication may need to address several distinct profiles rather than a single average respondent. For example, individuals with high knowledge but lower reported prevention may require messages focused on translating knowledge into action, whereas individuals with lower perceived relevance may require clearer communication about why VBDs are relevant in Switzerland. This is particularly important for mosquito-borne diseases, where individual and household practices such as removal of standing water depend on understanding both the vector ecology and the rationale for control.

### 4.3 Comparison with previous literature

This study is broadly consistent with previous European KAP studies on VBDs. The closest Swiss comparison is a study conducted in 2012 in Neuchâtel on preventive behaviours regarding Lyme disease [13]. In that study, 57% of the total population reported performing tick checks after outdoor activities, 53% reported wearing protective clothing, and 29% reported using tick repellents. These levels were lower than those observed in our Bern sample, where 79.3% reported tick checks, 70.8% protective clothing, and 51.3% repellent use. This difference may reflect regional variation, changes in public awareness or prevention messaging over the past decade, or differences in study populations. Interestingly, when Aenishaenslin et al. restricted their analysis to respondents familiar with Lyme disease, reported adoption of tick checks and protective clothing was higher than in the total Neuchâtel population, suggesting that disease awareness may be an important determinant of preventive behaviour.

The same pattern of uneven adoption across tick-prevention measures has been reported in other European settings. In the Netherlands, Beaujean et al. found relatively low adoption of preventive behaviours in the general population, ranging from 6% for repellent use to 37% for wearing protective clothing [11]. In Sweden, Slunge and Boman reported higher uptake of some measures, with 64% frequently using protective clothing, 63% performing tick checks, and 48% avoiding tall grass in areas with ticks; however, only 18% frequently tucked trousers into socks and 16% used repellents [12]. Our findings follow the same pattern, with tick checks and protective clothing reported more frequently than other preventive measures. Across studies, this suggests that simple personal behaviours are more readily adopted than measures perceived as restrictive, inconvenient, or less compatible with outdoor activities.

For mosquito-related prevention, the metropolitan French KAP survey provides a useful comparison [18]. In that study, the most frequently reported behaviours were storing containers out of the rain to avoid stagnant water (37%), applying skin repellent (36%), using mosquito candles or coils (35%), and removing stagnant water from flowerpots and vases (35%). In the Bern sample, individual protective behaviours were more frequently reported, including mosquito repellent use (66.5%) and window screens (42.6%). By contrast, removal of containers with standing water was reported by 29.6% of respondents. These results suggest that mosquito prevention in the Bern sample was more commonly oriented toward individual protection than environmental source reduction. Differences between the French and Swiss findings may reflect variation in perceived mosquito-borne disease risk, prevention campaigns, vector exposure, or norms around household-level control.

The association between tertiary university education and higher latent knowledge is also compatible with broader survey literature, including literature on selection and participation patterns in public health research [25, 26]. In our study, the sample was skewed toward higher educational attainment, which may have led to overestimation of population knowledge.

### 4.4 Strengths and limitations

This study has several strengths. It was embedded in an established cohort infrastructure, used a structured multi-stage assessment of knowledge, and incorporated internal validity checks through distractor items and contradiction screens. It also permitted direct comparison of tick- and mosquito-related domains within the same respondent population.

In addition, the analysis used an IRT model to derive a latent knowledge score while accounting for differences between items in difficulty and discrimination. This provided a more informative summary of knowledge than a count of correct responses. The clustering analysis also used a block-weighted Gower distance, which allowed binary and categorical KAP variables to be combined while preventing questionnaire domains with more items from dominating the cluster solution.

Several limitations should be considered. First, the survey was conducted within the canton of Bern and cannot be assumed to represent all of Switzerland. Regional differences in ecology, communication, and public experience with vectors may limit external validity. Second, participation was web-based and voluntary within a cohort framework, which likely contributed to selection toward more health-engaged and more highly educated participants. Third, the predefined validity filters improved interpretability of the KAP responses but may also have selected respondents with better comprehension of the questionnaire or greater engagement with the survey. Missingness was limited overall but concentrated in disease–vector matching items, and imputation was used for model-based analyses and clustering. Although this allowed complete-variable analyses, results may still be sensitive to the missing-data assumptions implicit in the imputation procedure.

Fourth, exposure and preventive behaviours were self-reported and therefore subject to recall and reporting biases. Fifth, the multivariable analysis was exploratory and should not be interpreted causally. Finally, the stakeholder survey was designed to provide contextual information and was not linked directly to population responses at the cantonal or municipal level.

### 4.5 Public health implications

The findings suggest several priorities for public health practice. First, communication on tick-borne diseases should be maintained, as it appears to be associated with solid public familiarity and widespread adoption of some protective behaviours. Second, communication on mosquito-borne diseases should be strengthened, with emphasis on diseases that are currently less well understood, including West Nile fever and chikungunya, and on why these risks are relevant to Switzerland even when local transmission remains limited or has not yet occurred.

Third, messaging should make stronger use of the public’s existing preference for practical, low-complexity actions. Household measures such as standing-water elimination may benefit from more explicit explanation and repeated communication.

Fourth, the association between tertiary university education and higher latent knowledge suggests that communication strategies should be designed for audiences with different levels of health literacy, prior scientific knowledge, and familiarity with public-health terminology.

Fifth, the contrast between stronger institutional engagement for mosquito-borne threats and stronger public familiarity with tick-borne risks suggests that preparedness planning should integrate risk communication more tightly with surveillance and control activities.

## 5 Conclusions

Among adults in the canton of Bern, knowledge and preventive practices regarding VBDs were stronger for tick-borne diseases than for mosquito-borne diseases. Ticks and tickborne diseases were more familiar and more clearly perceived as relevant in Switzerland, whereas mosquito bites were frequent but knowledge of mosquito-borne infections remained less complete. Exploratory multivariable analyses suggested that higher educational attainment and greater reported vector exposure were associated with higher knowledge scores, with domain-specific differences between tick- and mosquito-related knowledge. Complementary stakeholder findings indicate that institutional engagement is currently broader and more diversified for mosquito-borne diseases than for tick-borne diseases. This contrast between system-level activity and population-level understanding should be addressed through targeted communication and preparedness strategies. Strengthening mosquito-borne disease literacy while sustaining established tick prevention could improve readiness for both current and emerging VBD risks in Switzerland.

## Supporting information

All supplementary material

## Data Availability

Access to BEready data can be requested through the BEready data access procedure using the BEready data request form. Data access is subject to review and approval according to BEready governance and applicable ethical and legal requirements.
Anonymised stakeholder survey data may be made available upon reasonable request to Julien Riou, subject to approval by the Swiss National Centre for Climate Services (NCCS).

## Declarations

### Ethics approval and consent to participate

The general population (BEready) component of the study was approved by the responsible ethics committee (Kantonale Ethikkommission Bern) under Business Administration System for Ethics Committees (BASEC) numbers 2023-00333 and 2023-02290, and by the veterinary office of the canton of Bern (approval number BE21/2023). All BEready participants provided written informed consent prior to enrolment. The study is registered on ClinicalTrials.gov (NCT06739499).

The stakeholder survey collected professional information from cantonal and Liechtenstein authorities relating to their institutional role and activities, and did not involve the collection of personal health-related data. Participation in the stakeholder survey was voluntary, and respondents were informed of the purpose of the study before completing the questionnaire.

### Consent for publication

Not applicable.

### Availability of data and materials

Access to BEready data can be requested through the BEready data access procedure using the BEready data request form. Data access is subject to review and approval according to BEready governance and applicable ethical and legal requirements.

Anonymised stakeholder survey data may be made available upon reasonable request to Julien Riou, subject to approval by the Swiss National Centre for Climate Services (NCCS).

The analysis code used for this study is available on GitHub at: VBD_KAP_surveys.

### Competing interests

The authors declare that they have no competing interests.

### Funding

This work was supported by the Swiss National Centre for Climate Services (NCCS) through the NCCS-Impacts programme, which funded the doctoral position of LG and supported the design and conduct of the general population and stakeholder surveys. The BEready cohort infrastructure is supported by the University of Bern and the Multidisciplinary Center for Infectious Diseases. The funders had no role in the collection, analysis, or interpretation of the data, the writing of the manuscript, or the decision to submit it for publication.

### Authors’ contributions

LG conceived and designed the study, conducted the statistical analyses, and drafted the manuscript. JR helped in the design of the statistical analyses, contributed to data management, and provided overall supervision. AV secured funding for the study and provided overall supervision. EH helped in the design of the general population questionnaire and provided access to and guidance on the BEready cohort. AS and NH helped in the design and dissemination of the stakeholder survey. IM helped in the design of both questionnaires. All authors read, revised and approved the final manuscript.

## Acknowledgements

The authors thank the BEready participants and study team, and the cantonal and Liechtenstein stakeholders who completed the institutional survey.

## Notes

### Competing Interest Statement

The authors have declared no competing interest.

### Author Declarations

Ethics committee of University of Bern gave ethical approval for this work (Business Administration System for Ethics Committees (BASEC) numbers 2023-00333 and 2023-02290)

