## Supplementary material for "Knowledge, attitudes, practices regarding vector-borne diseases among adults in Switzerland: a cross-sectional survey": All supplementary material

### Supplementary Information for Knowledge, Attitudes and Practices on Vector-Borne Diseases in the Canton of Bern

#### S1.1 Sociodemography and exposure

##### Sociodemography

Table S1.1: Socio-demographic characteristics of respondents.

| **Characteristic** | **All respondents N = 1,847**[1] | **Analysis sample N = 1,337**[1] |
|---|---|---|
| **What sex were you assigned at birth?** | | |
| Female | 1,032 (55.9%) | 765 (57.2%) |
| Male | 815 (44.1%) | 572 (42.8%) |
| Intersex | 0 (0%) | 0 (0%) |
| **Which of the following labels best describes your social and felt gender?** | | |
| Female | 1,022 (55.8%) | 759 (57.2%) |
| Male | 804 (43.9%) | 566 (42.6%) |
| Other | 5 (0.27%) | 3 (0.23%) |
| (Missing) | 16 | 9 |
| **What is your highest completed education?** | | |
| None | 2 (0.11%) | 1 (0.08%) |
| Incomplete compulsory schooling | 2 (0.11%) | 0 (0%) |
| Compulsory schooling | 28 (1.58%) | 16 (1.24%) |
| One-year transitional program | 7 (0.40%) | 4 (0.31%) |
| General education school | 30 (1.69%) | 23 (1.78%) |
| Apprenticeship/vocational school | 343 (19.4%) | 228 (17.6%) |
| Gymnasial maturity | 93 (5.25%) | 69 (5.33%) |
| Professional maturity | 35 (1.98%) | 23 (1.78%) |
| Federal diploma exam | 180 (10.2%) | 127 (9.81%) |
| Higher vocational school | 208 (11.8%) | 153 (11.8%) |
| Bachelor degree | 261 (14.7%) | 187 (14.4%) |
| Master degree | 423 (23.9%) | 323 (24.9%) |
| Doctorate/habilitation | 158 (8.93%) | 141 (10.9%) |

*Continued on next page*

| Characteristic | All respondents N = 1,847[1] | Filtered sample N = 1,337[1] |
|---|---|---|
| (Missing) | 77 | 42 |
| **Since when have you been living in Switzerland?** | | |
| Since birth | 1,354 (73.5%) | 970 (72.7%) |
| Born in Switzerland but also lived abroad | 108 (5.87%) | 86 (6.45%) |
| Born abroad | 379 (20.6%) | 278 (20.8%) |
| (Missing) | 6 | 3 |
| **What is your marital status?** | | |
| Married, living with spouse | 970 (54.4%) | 714 (54.8%) |
| Married, permanently separated | 22 (1.23%) | 16 (1.23%) |
| Registered partnership, living together | 4 (0.22%) | 3 (0.23%) |
| Registered partnership, separated | 1 (0.06%) | 1 (0.08%) |
| Single, stable partnership | 292 (16.4%) | 230 (17.7%) |
| Single, living alone | 263 (14.8%) | 186 (14.3%) |
| Divorced | 172 (9.65%) | 119 (9.14%) |
| Widowed | 59 (3.31%) | 33 (2.53%) |
| (Missing) | 64 | 35 |
| **Occupation according to CH-ISCO 2019** | | |
| Administrative support staff | 356 (32.0%) | 287 (34.0%) |
| Directors, senior executives and managers | 8 (0.72%) | 6 (0.71%) |
| Intellectual and scientific professions | 121 (10.9%) | 84 (9.96%) |
| Intermediate occupations | 425 (38.2%) | 314 (37.2%) |
| Personal services staff, shopkeepers and sales assistants | 203 (18.2%) | 152 (18.0%) |
| (Missing) | 734 | 494 |

[1] n (%)

#### Exposures from inclusion questionnaire

Almost all participants (98.2%) reported having received vaccines at some point, and most indicated full mobility, with 87.1% having no problems walking and fewer than 4% reporting moderate to severe difficulties. A large majority (99.8%) had traveled abroad for more than two days, with Europe being the most frequently visited continent (94%), followed by North America (60.4%), Asia (54.1%), and Africa (46.5%).

[!t]

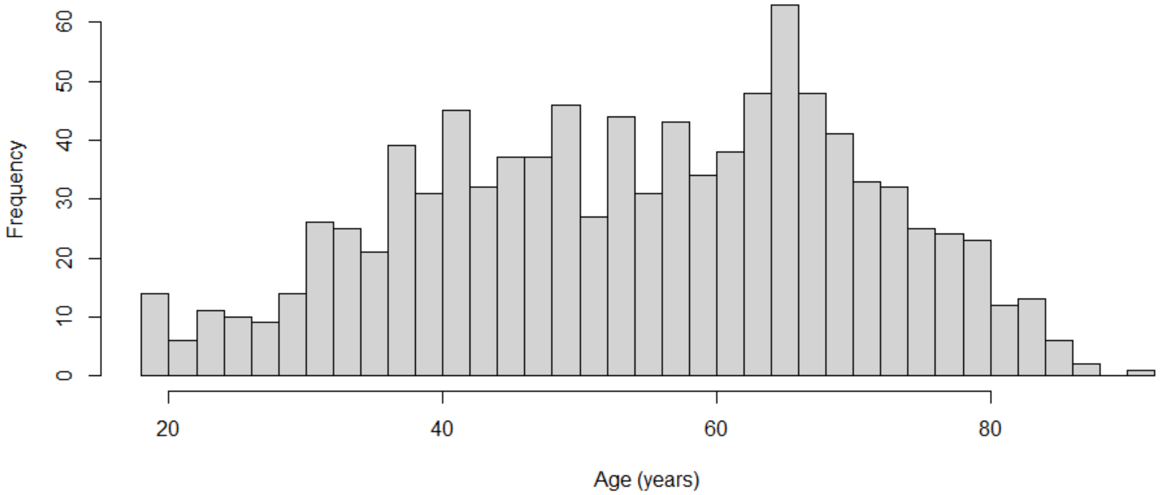


Figure S1.1: Age at inclusion

Table S1.2: Exposures from inclusion questionnaire.

| **Characteristic** | **All respondents N = 1,847**[1] | **Analysis sample N = 1,337**[1] |
|---|---|---|
| **Have you ever received any vaccines?** | | |
| Yes | 1,795 (98.5%) | 1,314 (98.7%) |
| No | 28 (1.54%) | 17 (1.28%) |
| (Missing) | 24 | 6 |
| **Please select what best describes your mobility today** | | |
| I have no problems walking around | 1,576 (86.8%) | 1,175 (87.9%) |
| I have slight problems walking around | 171 (9.42%) | 115 (8.61%) |
| I have moderate problems walking around | 51 (2.81%) | 35 (2.62%) |
| I have severe problems walking around | 18 (0.99%) | 11 (0.82%) |
| I am unable to walk around | 0 (0%) | 0 (0%) |
| (Missing) | 31 | 1 |
| **Have you ever stayed abroad for more than two days?** | | |
| Yes | 1,800 (99.8%) | 1,331 (99.8%) |
| No | 4 (0.22%) | 2 (0.15%) |
| (Missing) | 43 | 4 |
| **On which continent?** | | |
| Europe | 1,763 (95.5%) | 1,307 (97.8%) |

*Continued on next page*

| Characteristic | All respondents N = 1,847[1] | Analysis sample N = 1,337[1] |
|---|---|---|
| North America | 1,149 (62.2%) | 867 (64.8%) |
| Central America | 361 (19.5%) | 277 (20.7%) |
| South America | 448 (24.3%) | 346 (25.9%) |
| Africa | 885 (47.9%) | 676 (50.6%) |
| Asia | 1,007 (54.5%) | 764 (57.1%) |
| Oceania | 404 (21.9%) | 309 (23.1%) |
| (Missing) | 0 | 0 |

[1] n (%)

The EQ VAS records the patient's self-rated health on a vertical visual analogue scale where the endpoints are labelled "The best health you can imagine" and "The worst health you can imagine". The VAS can be used as a quantitative measure of health outcome that reflects the patient's own judgement. We observe an average of 83 (SD 13.2), consistent with a representative survey in French-speaking Switzerland reporting a mean EQ-VAS of 81.7 (SD 15.5) [1].

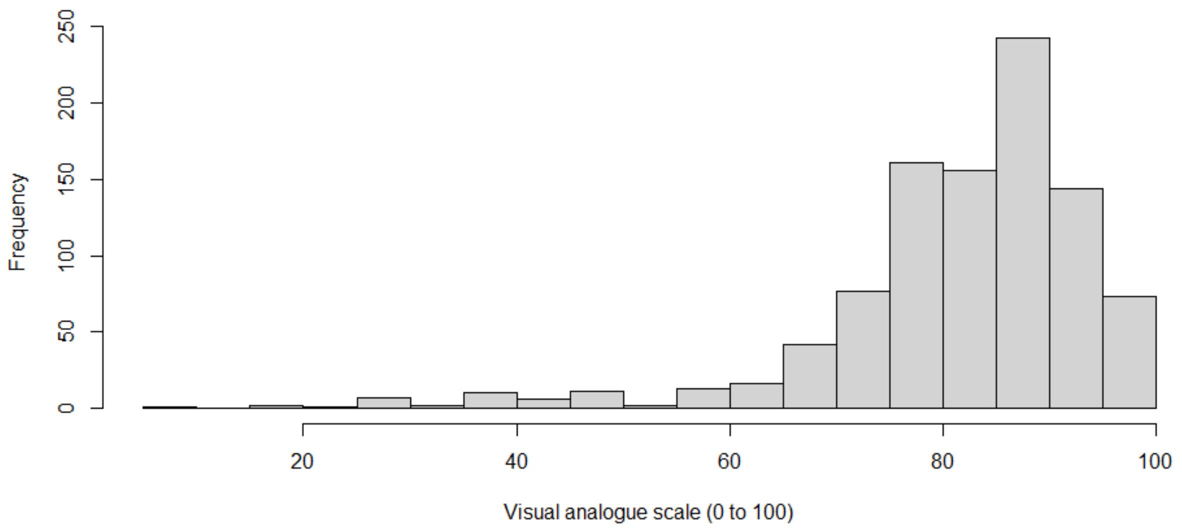


Figure S1.2: EQ-VAS scale

#### Municipality of residence

For each municipality in the canton of Bern, we mapped the number of survey respondents. The map illustrates the geographic distribution of the surveyed population, with a markedly higher concentration of respondents in the city of Bern than in the rest of the canton.

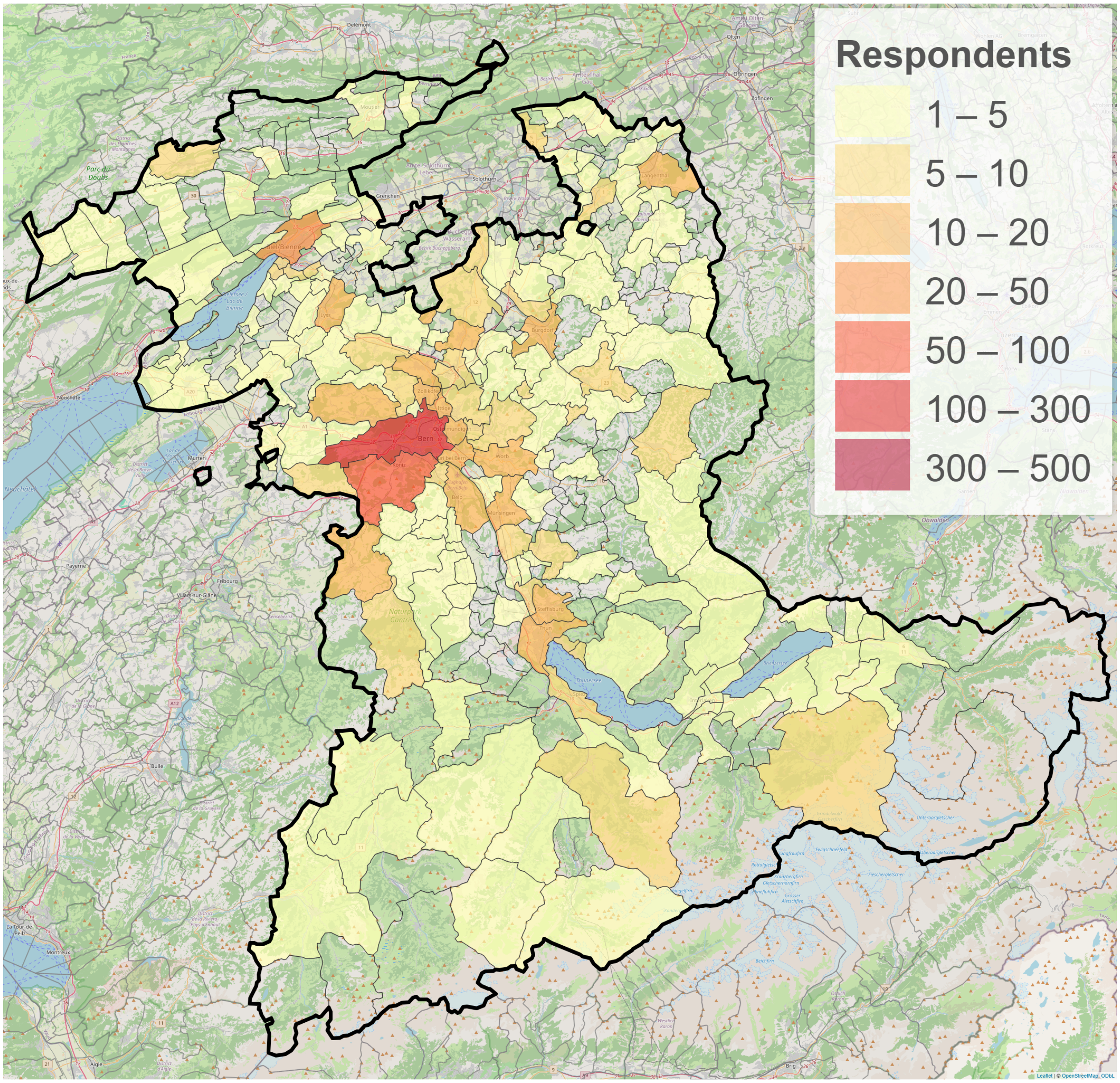


Figure S1.3: Municipality of residence of survey respondents.

#### S1.2 General knowledge

##### Baseline control variables

Table S1.3: Understanding check up.

| **Introductory text** | **All respondents N = 1,847**[1] | **Analysis sample N = 1,337**[1] |
|---|---|---|
| **Please read the following text carefully before answering the questions: A vector is an organism that can transmit a pathogen (virus, bacterium, or parasite), primarily by feeding on an infected host (human or animal), and then transmitting it to other hosts. Tick-borne encephalitis is a disease transmitted by tick bites. Would you classify it as a vector-borne disease?** | | |
| Yes | 1,415 (78.4%) | 1,337 (100.0%) |
| No | 201 (11.1%) | 0 (0%) |
| I don't know | 189 (10.5%) | 0 (0%) |
| (Missing) | 42 | 0 |

[1] n (%)

Table S1.4: Preliminary knowledge check-up.

| **Question** | **All respondents N = 1,847**[1] | **Analysis sample N = 1,337**[1] |
|---|---|---|
| **Have you heard of the following diseases?** | | |
| West Nile fever | 594 (32.2%) | 510 (38.1%) |
| Dengue | 1,634 (88.5%) | 1,243 (93.0%) |
| Zika | 1,176 (63.7%) | 967 (72.3%) |
| Chikungunya | 426 (23.1%) | 373 (27.9%) |
| Lyme disease | 1,416 (76.7%) | 1,123 (84.0%) |
| Influenza | 1,751 (94.8%) | 1,309 (97.9%) |
| Measles | 1,770 (95.8%) | 1,319 (98.7%) |
| None of these | 5 (0.27%) | 1 (0.07%) |
| (Missing) | 0 | 0 |
| **Have you heard of the following organisms?** | | |
| Ticks | 1,791 (97.0%) | 1,331 (99.6%) |
| Mosquitoes | 1,783 (96.5%) | 1,328 (99.3%) |
| Wasps | 1,731 (93.7%) | 1,295 (96.9%) |
| Bed bugs | 1,716 (92.9%) | 1,285 (96.1%) |
| None of these | 4 (0.22%) | 0 (0%) |
| (Missing) | 0 | 0 |

[1] n (%)

A majority of participants answered consistently when linking vectors to disease transmission, but a minority displayed contradictions. About 6% denied ticks or mosquitoes as transmitters while simultaneously identifying them as responsible for disease transmission. In total, 10.7% of respondents contradicted themselves in at least one case. This suggests that while knowledge was generally coherent, a small but notable share of respondents held conflicting beliefs or had difficulties applying knowledge consistently.

Table S1.5: Logical contradictions in respondents' answers.

| **Question** | **All respondents N = 1,847**[1] | **Analysis sample N = 1,337**[1] |
|---|---|---|
| **Denied tick as transmitter and identified ticks as transmiting a disease** | | |
| Consistent | 1,702 (94.6%) | 1,337 (100.0%) |
| Contradiction | 97 (5.39%) | 0 (0%) |
| (Missing) | 48 | 0 |
| **Denied mosquitoes transmitter and identified mosquitoes as transmiting a disease** | | |
| Consistent | 1,676 (93.7%) | 1,337 (100.0%) |
| Contradiction | 113 (6.32%) | 0 (0%) |
| (Missing) | 58 | 0 |
| **Any of the two contradiction** | | |
| Consistent | 1,604 (89.8%) | 1,337 (100.0%) |
| Contradiction | 182 (10.2%) | 0 (0%) |
| (Missing) | 61 | 0 |

[1] n (%)

#### Filtering

To ensure data quality and consistency, we used Supplementary Tables S1.3 and S1.5 to exclude respondents who either lacked a basic understanding of vector-borne diseases or provided logically inconsistent answers throughout the questionnaire. This process ensures that responses reflect a reliable snapshot of participants' knowledge and that the respondents do not adapt their answers during the completion of the questionnaire. In total, 282 individuals were excluded based on these criteria, leaving a final sample of 709 participants for analyzing the results of follow-up questions concerning knowledge on vector-borne diseases.

#### Item-level accuracy

Table S1.6: Vector-borne disease test item.

| **Test item** | **All respondents N = 1,847**[1] | **Analysis sample N = 1,337**[1] |
|---|---|---|
| **Question : Which of these diseases do you think are vector-borne diseases?** | | |
| (Missing) | 0 | 0 |
| **West Nile correctly classified as VBD** | | |
| Incorrect | 1,157 (62.6%) | 760 (56.8%) |
| Correct | 690 (37.4%) | 577 (43.2%) |
| **Dengue correctly classified as VBD** | | |
| Incorrect | 418 (22.6%) | 173 (12.9%) |
| Correct | 1,429 (77.4%) | 1,164 (87.1%) |
| **Zika correctly classified as VBD** | | |
| Incorrect | 723 (39.1%) | 398 (29.8%) |
| Correct | 1,124 (60.9%) | 939 (70.2%) |
| **Chikungunya correctly classified as VBD** | | |
| Incorrect | 1,293 (70.0%) | 862 (64.5%) |
| Correct | 554 (30.0%) | 475 (35.5%) |
| **Lyme correctly classified as VBD** | | |
| Incorrect | 551 (29.8%) | 222 (16.6%) |
| Correct | 1,296 (70.2%) | 1,115 (83.4%) |
| **Influenza correctly classified as not a VBD** | | |
| Incorrect | 427 (23.1%) | 261 (19.5%) |
| Correct | 1,420 (76.9%) | 1,076 (80.5%) |
| **Measles correctly classified as not a VBD** | | |
| Incorrect | 356 (19.3%) | 212 (15.9%) |
| Correct | 1,491 (80.7%) | 1,125 (84.1%) |

[1] n (%)

Table S1.7: Vector test item.

| **Test item** | **All respondents N = 1,847**[1] | **Analysis sample N = 1,337**[1] |
|---|---|---|
| **Question : Which of these organisms transmit diseases to humans?** | | |
| (Missing) | 0 | 0 |
| **Ticks correctly identified as transmitting disease** | | |
| Incorrect | 169 (9.15%) | 1 (0.07%) |

*Continued on next page*

| Test item | All respondents N = 1,847[1] | Analysis sample N = 1,337[1] |
|---|---|---|
| Correct | 1,678 (90.9%) | 1,336 (99.9%) |
| **Mosquitoes correctly identified as transmitting disease** | | |
| Incorrect | 224 (12.1%) | 18 (1.35%) |
| Correct | 1,623 (87.9%) | 1,319 (98.7%) |
| **Wasps correctly identified as not transmitting disease** | | |
| Incorrect | 109 (5.90%) | 73 (5.46%) |
| Correct | 1,738 (94.1%) | 1,264 (94.5%) |
| **Bed bugs identified as not transmitting disease** | | |
| Incorrect | 376 (20.4%) | 285 (21.3%) |
| Correct | 1,471 (79.6%) | 1,052 (78.7%) |

[1] n (%)

Table S1.8: Disease–vector matching test item.

| Test item | All respondents N = 1,847[1] | Analysis sample N = 1,337[1] |
|---|---|---|
| **Question : Match each disease on the left with its vector on the right (one per row). E.g. 'tick-borne encephalitis' -> 'ticks'.** | | |
| **TBE correctly matched to ticks** | | |
| Incorrect | 61 (3.41%) | 20 (1.50%) |
| Correct | 1,726 (96.6%) | 1,310 (98.5%) |
| **West Nile correctly matched to mosquitoes** | | |
| Incorrect | 708 (43.3%) | 489 (40.1%) |
| Correct | 927 (56.7%) | 731 (59.9%) |
| **Dengue correctly matched to mosquitoes** | | |
| Incorrect | 276 (15.7%) | 165 (12.5%) |
| Correct | 1,483 (84.3%) | 1,150 (87.5%) |
| **Zika correctly matched to mosquitoes** | | |
| Incorrect | 576 (34.3%) | 354 (28.1%) |
| Correct | 1,101 (65.7%) | 906 (71.9%) |
| **Chikungunya correctly matched to mosquitoes** | | |
| Incorrect | 940 (59.3%) | 654 (55.3%) |
| Correct | 645 (40.7%) | 529 (44.7%) |
| **Lyme correctly matched to ticks** | | |
| Incorrect | 300 (17.0%) | 161 (12.2%) |

*Continued on next page*

| Test item | All respondents N = 1,847[1] | Analysis sample N = 1,337[1] |
|---|---|---|
| Correct | 1,461 (83.0%) | 1,159 (87.8%) |
| **Influenza correctly matched to none** | | |
| Incorrect | 177 (10.8%) | 93 (7.74%) |
| Correct | 1,456 (89.2%) | 1,109 (92.3%) |
| **Measles correctly matched to none** | | |
| Incorrect | 214 (13.1%) | 113 (9.42%) |
| Correct | 1,414 (86.9%) | 1,086 (90.6%) |

[1] n (%)

### S1.3 Individual exposure and prevention

#### Individual risk exposure

Table S1.9: Self-reported individual exposure.

| Question | All respondents N = 1,847[1] | Analysis sample N = 1,337[1] |
|---|---|---|
| **How many tick bites do you receive on average per year (spring to autumn)?** | | |
| 0 | 1,090 (60.6%) | 784 (58.9%) |
| 1–3 | 616 (34.2%) | 471 (35.4%) |
| 4–6 | 49 (2.72%) | 38 (2.85%) |
| 7–9 | 19 (1.06%) | 15 (1.13%) |
| 10+ | 26 (1.44%) | 24 (1.80%) |
| (Missing) | 47 | 5 |
| **In summer, how often are you bitten by mosquitoes?** | | |
| Every day | 157 (8.91%) | 119 (9.08%) |
| At least weekly | 758 (43.0%) | 573 (43.7%) |
| At least monthly | 530 (30.1%) | 393 (30.0%) |
| At least yearly | 164 (9.31%) | 122 (9.31%) |
| Less than yearly | 42 (2.38%) | 30 (2.29%) |
| Never | 27 (1.53%) | 20 (1.53%) |
| I don’t know | 84 (4.77%) | 53 (4.05%) |
| (Missing) | 85 | 27 |

[1] n (%)

#### Individual prevention measures

Table S1.10: Self-reported preventive practices.

| Question | All respondents N = 1,847[1] | Analysis sample N = 1,337[1] |
|---|---|---|
| **In the last 12 months, what have you done to protect yourself/family against tick bites or tick-borne diseases?** | | |
| Check for and remove ticks after outdoor activities | 1,379 (74.7%) | 1,060 (79.3%) |
| Wear protective clothing | 1,242 (67.2%) | 947 (70.8%) |
| Use insect repellent on skin or clothing | 893 (48.3%) | 686 (51.3%) |
| Avoid wooded areas during tick activity | 221 (12.0%) | 161 (12.0%) |
| Other measures (open-text) | 243 (13.2%) | 183 (13.7%) |
| No active measures | 163 (8.83%) | 107 (8.00%) |
| (Missing) | 0 | 0 |
| **In the last 12 months, what have you done to protect yourself/family against mosquito bites?** | | |
| Use repellent | 1,183 (64.0%) | 889 (66.5%) |
| Wear protective clothing | 799 (43.3%) | 612 (45.8%) |
| Burn mosquito coils | 296 (16.0%) | 234 (17.5%) |
| Stay in screened areas | 319 (17.3%) | 255 (19.1%) |
| Install window screens | 780 (42.2%) | 569 (42.6%) |
| Use fans | 127 (6.88%) | 95 (7.11%) |
| Use automatic insect spray | 151 (8.18%) | 114 (8.53%) |
| Install bed nets | 257 (13.9%) | 192 (14.4%) |
| Remove containers with standing water | 507 (27.4%) | 396 (29.6%) |
| Other measures (open-text) | 99 (5.36%) | 79 (5.91%) |
| No active measures | 207 (11.2%) | 138 (10.3%) |
| (Missing) | 0 | 0 |

[1] n (%)

#### Individual prevention measures: open-ended answers

Most respondents reported vaccination against tick-borne encephalitis (FSME/TBE) as their main preventive measure, often mentioning it explicitly for themselves or their children. A smaller number referred to protecting pets, for example with tick collars or treatments, as an indirect preventive strategy. Few participants mentioned other measures such as avoiding tall grass or forests, wearing protective clothing or hats, using repellents, including sprays, oils, or patches, or employing unconventional approaches such as vinegar, vitamin B, or ceramic bracelets. Overall, vaccination was by far the most commonly cited individual preventive action, with other measures appearing rarely and in isolated cases.

Respondents described a wide range of individual strategies to prevent mosquito bites. Common measures included the use of repellents such as insect sprays, mosquito-repellent candles, incense sticks, essential oils, for example lavender, clove, or oregano, and specialized diffusers or bracelets. Several participants reported installing or using physical barriers such as mosquito nets, insect screens, or curtains, and others emphasized behavioral measures, including keeping lights off in the evening, ventilating rooms during the day, and avoiding open windows at night. Some mentioned directly eliminating mosquitoes indoors through catching, swatting, or using electric devices. Additional approaches included reducing breeding sites by covering or regularly changing standing water, maintaining bat-friendly gardens, or applying larvicides. A few respondents also referred to vaccination or the use of vitamin supplements as protective measures.

Open-ended answers can be requested upon reasonable request to the corresponding author.

#### S1.4 Mitigation strategies, vector control, and perceived relevance

##### Mitigation strategies and vector control

Table S1.11: Beliefs regarding effectiveness of public control measures.

| **Question** | **All respondents** N = 1,847[1] | **Analysis sample** N = 1,337[1] |
|---|---|---|
| **Which of these do you think effectively protect the public against tick-borne diseases?** | | |
| Apply environmental pesticides | 60 (3.25%) | 44 (3.29%) |
| Use biological control (e.g. predators) | 730 (39.5%) | 572 (42.8%) |
| Clear vegetation in woods | 31 (1.68%) | 25 (1.87%) |
| Protect deer from ticks | 126 (6.82%) | 94 (7.03%) |
| Control deer numbers in public woods | 65 (3.52%) | 46 (3.44%) |
| Block deer from public woods | 20 (1.08%) | 13 (0.97%) |
| Protect small rodents from ticks | 94 (5.09%) | 69 (5.16%) |
| Promote personal protective measures | 1,463 (79.2%) | 1,146 (85.7%) |
| None of these | 161 (8.72%) | 90 (6.73%) |

*Continued on next page*

| Question | All respondents N = 1,847[1] | Analysis sample N = 1,337[1] |
|---|---|---|
| (Missing) | 0 | 0 |
| **Which of these do you think effectively protect the public against mosquito-borne diseases?** | | |
| Apply environmental pesticides | 88 (4.76%) | 61 (4.56%) |
| Use biological control (e.g. predators) | 754 (40.8%) | 597 (44.7%) |
| Eliminate standing water in containers | 1,098 (59.4%) | 878 (65.7%) |
| Apply larvicides | 222 (12.0%) | 187 (14.0%) |
| Install repellent sprayers | 204 (11.0%) | 152 (11.4%) |
| Install mosquito traps | 652 (35.3%) | 500 (37.4%) |
| Release irradiated or genetically modified mosquitoes | 363 (19.7%) | 308 (23.0%) |
| Promote personal protective measures | 1,420 (76.9%) | 1,112 (83.2%) |
| None of these | 82 (4.44%) | 45 (3.37%) |
| (Missing) | 0 | 0 |

[1] n (%)

### Perceived relevance of VBDs

Table S1.12: Perception on vector-borne disease risk and control measures.

| Question | All respondents N = 1,847[1] | Analysis sample N = 1,337[1] |
|---|---|---|
| **Do you consider the following disease as a current health problem in Switzerland?** | | |
| Tick-borne encephalitis | 1,353 (73.3%) | 1,044 (78.1%) |
| West Nile fever | 48 (2.60%) | 41 (3.07%) |
| Dengue | 165 (8.93%) | 115 (8.60%) |
| Zika | 108 (5.85%) | 83 (6.21%) |
| Chikungunya | 43 (2.33%) | 34 (2.54%) |
| Lyme disease | 1,163 (63.0%) | 933 (69.8%) |
| (Missing) | 0 | 0 |
| **Do you think vector control measures are necessary in Switzerland?** | | |
| Yes | 862 (47.9%) | 662 (49.7%) |
| No | 349 (19.4%) | 270 (20.3%) |
| Don't know | 588 (32.7%) | 401 (30.1%) |
| (Missing) | 48 | 4 |

[1] n (%)

### Supplementary Information for Cantonal Engagement in the Prevention and Control of Vector-Borne Diseases in Switzerland and Liechtenstein

#### S2.1 Involvement and resources

Table S2.1: Involvement and resources in the field of mosquito- and tick-borne diseases.

| **Question** | **Overall** N = 55[1] | **Human health** N = 22[1] | **Animal health** N = 17[1] | **Environment** N = 16[1] |
|---|---|---|---|---|
| **Is your authority currently involved in the implementation of measures or activities (e.g., public relations, monitoring, or control) in the area of diseases transmitted by ticks and mosquitoes?** | | | | |
| Yes | 41 (77%) | 14 (67%) | 16 (94%) | 11 (73%) |
| No | 12 (23%) | 7 (33%) | 1 (6%) | 4 (27%) |
| No information / don't know | 0 | 0 | 0 | 0 |
| (Missing) | 2 | 1 | 0 | 1 |
| **If you answered no, what are the reasons?** | | | | |
| The introduction of measures/activities in the area of diseases transmitted by ticks and mosquitoes is currently being reviewed/planned. | 0 | 0 | 0 | 0 |
| Diseases transmitted by ticks and mosquitoes are not considered part of the responsibilities of your authority. | 5 (42%) | 1 (14%) | 0 | 4 (100%) |
| Diseases transmitted by ticks and mosquitoes are not a high priority for your authority. | 1 (8%) | 1 (14%) | 0 | 0 |
| The number of cases of diseases transmitted by ticks and mosquitoes is not yet a significant concern to trigger measures in my area. | 2 (17%) | 2 (29%) | 0 | 0 |
| Lack of financial and/or human resources. | 6 (50%) | 5 (71%) | 1 (100%) | 0 |
| Lack of professionally trained personnel. | 1 (8%) | 1 (14%) | 0 | 0 |
| Lack of public interest. | 0 | 0 | 0 | 0 |
| Lack of political decision/mandate. | 1 (8%) | 1 (14%) | 0 | 0 |
| Lack of national guidelines/basics/recommendations to follow. | 1 (8%) | 1 (14%) | 0 | 0 |
| No information / don't know | 2 (17%) | 2 (29%) | 0 | 0 |
| Other reasons | 0 | 0 | 0 | 0 |
| **Do you know of any strategy or action plan in your canton regarding diseases transmitted by ticks and mosquitoes?** | | | | |
| Yes, for tick-borne diseases | 6 (11%) | 3 (14%) | 3 (18%) | 0 |

*Continued on next page*

| Question | Overall N = 55[1] | Human health N = 22[1] | Animal health N = 17[1] | Environment N = 16[1] |
|---|---|---|---|---|
| Yes, for mosquito-borne diseases | 27 (49%) | 8 (36%) | 11 (65%) | 8 (50%) |
| No, neither for tick-borne nor mosquito-borne diseases | 18 (33%) | 11 (50%) | 4 (24%) | 3 (19%) |
| No information / don't know | 11 (20%) | 3 (14%) | 2 (12%) | 6 (38%) |
| (Missing) | 0 | | | |
| **Does your authority have human resources to develop and implement activities related to diseases transmitted by ticks and mosquitoes?** | | | | |
| Yes, for tick-borne diseases | 6 (11%) | 5 (23%) | 1 (6%) | 0 |
| Yes, for mosquito-borne diseases | 22 (40%) | 9 (41%) | 5 (29%) | 8 (50%) |
| No, neither for tick-borne nor mosquito-borne diseases | 27 (49%) | 12 (55%) | 11 (65%) | 4 (25%) |
| No information / don't know | 6 (11%) | 1 (5%) | 1 (6%) | 4 (25%) |
| (Missing) | 0 | | | |
| **Does your authority have a budget for the development and implementation of activities in the field of diseases transmitted by ticks and mosquitoes?** | | | | |
| Yes, for tick-borne diseases | 4 (7%) | 2 (9%) | 1 (6%) | 1 (6%) |
| Yes, for mosquito-borne diseases | 18 (33%) | 9 (41%) | 2 (12%) | 7 (44%) |
| No, neither for tick-borne nor mosquito-borne diseases | 30 (55%) | 11 (50%) | 13 (76%) | 6 (38%) |
| No information / don't know | 6 (11%) | 2 (9%) | 2 (12%) | 2 (13%) |
| (Missing) | 0 | | | |

[1] n (%)

#### S2.2 Concrete measures and activities

Table S2.2: Concrete measures and activities in the field of mosquito- and tick-borne diseases (part 1).

| Question | Overall N = 55[1] | Human health N = 22[1] | Animal health N = 17[1] | Environment N = 16[1] |
|---|---|---|---|---|
| **What activities is your authority currently (2024/2025) implementing in the field of tick-borne diseases?** | | | | |
| Public relations (distribution of educational material about tick-borne diseases to the general public) | 12 (22%) | 9 (41%) | 3 (18%) | 0 |
| Targeted awareness of professionals (documents on tick-borne diseases) | 7 (13%) | 6 (27%) | 1 (6%) | 0 |
| Vaccination advice for the population (by health personnel) | 3 (5%) | 3 (14%) | 0 | 0 |

*Continued on next page*

| Question | Overall N = 55[1] | Human health N = 22[1] | Animal health N = 17[1] | Environment N = 16[1] |
|---|---|---|---|---|
| Advice for homeowners on tick management on their property | 0 | 0 | 0 | 0 |
| Active tick monitoring (collecting ticks in the field for identification, examination, or analysis) | 0 | 0 | 0 | 0 |
| Passive tick monitoring (acceptance of tick samples submitted by the public, veterinarians, doctors, etc., for identification, examination, or analysis) | 2 (4%) | 1 (5%) | 1 (6%) | 0 |
| Training and distribution of educational materials | 0 | 0 | 0 | 0 |
| No information / don't know | 6 (11%) | 3 (14%) | 0 | 3 (19%) |
| Other: open answer | 10 (18%) | 5 (23%) | 5 (29%) | 0 |
| My authority does not conduct any activities for tick-borne diseases | 29 (53%) | 5 (23%) | 11 (65%) | 13 (81%) |
| (Missing) | 0 | | | |
| **What activities is your authority currently (2024/2025) implementing in the field of mosquito-borne diseases?** | | | | |
| Public relations (distribution of educational material about mosquito-borne diseases to the general public) | 22 (40%) | 5 (23%) | 9 (53%) | 8 (50%) |
| Targeted awareness of professionals (documents on mosquito-borne diseases) | 22 (40%) | 6 (27%) | 12 (71%) | 4 (25%) |
| Active mosquito monitoring (collecting mosquitoes using traps for identification and diagnostics, e.g., Ovitraps) | 20 (36%) | 7 (32%) | 0 | 13 (81%) |
| Passive mosquito monitoring (acceptance of mosquito submissions and sightings directly from the public or via the Swiss Mosquito Network) | 18 (33%) | 5 (23%) | 0 | 13 (81%) |
| Verification of mosquito sightings at new locations | 16 (29%) | 4 (18%) | 0 | 12 (75%) |
| Mosquito control on public land | 10 (18%) | 5 (23%) | 1 (6%) | 4 (25%) |
| Mosquito control on private properties | 8 (15%) | 3 (14%) | 3 (18%) | 2 (13%) |
| Training and distribution of educational materials | 11 (20%) | 4 (18%) | 2 (12%) | 5 (31%) |
| No information / don't know | 1 (2%) | 1 (5%) | 0 | 0 |
| Other: open answer | 17 (31%) | 6 (27%) | 6 (35%) | 5 (31%) |
| My authority does not conduct any activities for mosquito-borne diseases | 11 (20%) | 8 (36%) | 2 (12%) | 1 (6%) |
| (Missing) | 0 | | | |
| **If your authority develops information, what subjects are covered?** | | | | |
| Correct removal of ticks | 5 (29%) | 5 (45%) | 0 | 0 |
| Elimination of mosquito breeding sites | 8 (47%) | 4 (36%) | 4 (67%) | 0 |
| Vaccinations | 11 (65%) | 7 (64%) | 4 (67%) | 0 |
| Information on mosquito/tick sprays | 5 (29%) | 5 (45%) | 0 | 0 |

*Continued on next page*

| Question | Overall N = 55[1] | Human health N = 22[1] | Animal health N = 17[1] | Environment N = 16[1] |
|---|---|---|---|---|
| Protection against bites/stings | 9 (53%) | 7 (64%) | 2 (33%) | 0 |
| Awareness for careful symptom monitoring | 9 (53%) | 6 (55%) | 3 (50%) | 0 |
| Information on risk areas | 8 (47%) | 6 (55%) | 2 (33%) | 0 |
| Information on the correct installation of mosquito nets | 0 | 0 | 0 | 0 |
| No information / don't know | 2 (12%) | 2 (18%) | 0 | 0 |
| Other: open answer | 2 (12%) | 1 (9%) | 1 (17%) | 0 |
| (Missing) | 38 | | | |

[1] n (%)

Table S2.3: Concrete measures and activities in the field of mosquito- and tick-borne diseases (part 2).

| Question | Overall N = 55[1] | Human health N = 22[1] | Animal health N = 17[1] | Environment N = 16[1] |
|---|---|---|---|---|
| **Among the following fields of action, where is the greatest need for information or the greatest lack of knowledge about tick-borne diseases in the authority you work for?** | | | | |
| Prevention of tick-borne diseases | 10 (18%) | 7 (32%) | 3 (18%) | 0 |
| Detection of ticks and pathogens in ticks | 10 (18%) | 6 (27%) | 4 (24%) | 0 |
| Diagnosis of tick-borne diseases | 3 (5%) | 3 (14%) | 0 | 0 |
| Management and control of tick-borne diseases | 11 (20%) | 7 (32%) | 2 (12%) | 2 (13%) |
| Therapy/treatment of tick-borne diseases | 4 (7%) | 2 (9%) | 2 (12%) | 0 |
| Other: open answer | 15 (27%) | 3 (14%) | 7 (41%) | 5 (31%) |
| (Missing) | 0 | | | |
| **Among the following fields of action, where is the greatest need for information or the greatest lack of knowledge about mosquito-borne diseases in the authority you work for?** | | | | |
| Prevention of mosquito-borne diseases | 12 (22%) | 6 (27%) | 4 (24%) | 2 (13%) |
| Detection of invasive mosquitoes and pathogens in mosquitoes | 11 (20%) | 8 (36%) | 3 (18%) | 0 |
| Diagnosis of mosquito-borne diseases | 6 (11%) | 3 (14%) | 1 (6%) | 2 (13%) |
| Management and control of mosquitoes/mosquito-borne diseases | 20 (36%) | 11 (50%) | 5 (29%) | 4 (25%) |
| Therapy/treatment of mosquito-borne diseases | 6 (11%) | 3 (14%) | 2 (12%) | 1 (6%) |
| Other: open answer | 15 (27%) | 6 (27%) | 4 (24%) | 5 (31%) |
| (Missing) | 0 | | | |

*Continued on next page*

| Question | Overall N = 55[1] | Human health N = 22[1] | Animal health N = 17[1] | Environment N = 16[1] |
|---|---|---|---|---|
| **What measures is your authority implementing to control or combat invasive mosquitoes?** | | | | |
| Adulticides (Pyrethroids) | 6 (11%) | 3 (14%) | 2 (12%) | 1 (6%) |
| Non-chemical larvicides | 4 (7%) | 2 (9%) | 0 | 2 (13%) |
| Biological larvicides | 14 (25%) | 5 (23%) | 0 | 9 (56%) |
| Elimination of breeding sites | 11 (20%) | 4 (18%) | 0 | 7 (44%) |
| Sterilization of mosquitoes | 1 (2%) | 1 (5%) | 0 | 0 |
| None | 29 (53%) | 14 (64%) | 10 (59%) | 5 (31%) |
| Other: open answer | 19 (35%) | 4 (18%) | 9 (53%) | 6 (38%) |
| (Missing) | 0 | | | |

[1] n (%)

#### S2.3 Collaboration and coordination

Table S2.4: Collaboration and coordination in the field of mosquito- and tick-borne diseases.

| Question | Overall N = 55[1] | Human health N = 22[1] | Animal health N = 17[1] | Environment N = 16[1] |
|---|---|---|---|---|
| **Does your authority maintain an active network or exchanges with researchers and specialists in the field?** | | | | |
| Yes, for tick-borne diseases | 11 (20%) | 5 (23%) | 6 (35%) | 0 |
| Yes, for mosquito-borne diseases | 33 (60%) | 12 (55%) | 10 (59%) | 11 (69%) |
| No, neither for tick-borne nor mosquito-borne diseases | 15 (27%) | 7 (32%) | 5 (29%) | 3 (19%) |
| No information / don't know | 7 (13%) | 3 (14%) | 2 (12%) | 2 (13%) |
| (Missing) | 0 | | | |
| **Is the issue of tick- and mosquito-borne diseases sufficiently or inadequately addressed in the implementation of the national climate change adaptation strategy?** | | | | |
| Yes, sufficient for tick-borne diseases | 9 (16%) | 5 (23%) | 3 (18%) | 1 (6%) |
| Yes, sufficient for mosquito-borne diseases | 11 (20%) | 5 (23%) | 3 (18%) | 3 (19%) |
| Insufficient for tick-borne diseases | 4 (7%) | 2 (9%) | 1 (6%) | 1 (6%) |
| Insufficient for mosquito-borne diseases | 3 (5%) | 2 (9%) | 1 (6%) | 0 |
| No, neither for tick-borne nor mosquito-borne diseases | 6 (11%) | 2 (9%) | 4 (24%) | 0 |
| No information / don't know | 36 (65%) | 13 (59%) | 10 (59%) | 13 (81%) |

*Continued on next page*

| Question | Overall N = 55[1] | Human health N = 22[1] | Animal health N = 17[1] | Environment N = 16[1] |
|---|---|---|---|---|
| (Missing) | 0 | | | |
| **Is the issue of diseases transmitted by ticks and mosquitoes sufficiently or insufficiently taken into account in the implementation of your canton's climate change adaptation strategy (if any)?** | | | | |
| Yes, sufficient for tick-borne diseases | 10 (18%) | 5 (23%) | 4 (24%) | 1 (6%) |
| Yes, sufficient for mosquito-borne diseases | 19 (35%) | 7 (32%) | 4 (24%) | 8 (50%) |
| Insufficient for tick-borne diseases | 6 (11%) | 2 (9%) | 2 (12%) | 2 (13%) |
| Insufficient for mosquito-borne diseases | 3 (5%) | 1 (5%) | 2 (12%) | 0 |
| No, neither for tick-borne nor mosquito-borne diseases | 4 (7%) | 2 (9%) | 2 (12%) | 0 |
| No information / don't know | 29 (53%) | 12 (55%) | 9 (53%) | 8 (50%) |
| (Missing) | 0 | | | |

[1] n (%)

#### S2.4 Open-text answers

Open-ended answers can be requested upon reasonnable request to the corresponding author.

#### References


[1] Perneger TV, Combescure C, Courvoisier DS. General Population Reference Values for the French Version of the EuroQol EQ-5D Health Utility Instrument;13(5):631-5. Available from: https://www.sciencedirect.com/science/article/pii/S1098301510601052.
